# Socio-economic inequalities in adolescent mental health in the UK: multiple socio-economic indicators and reporter effects

**DOI:** 10.1101/2022.01.13.22269209

**Authors:** Matthew Hazell, Emma Thornton, Hassan Haghparast-Bidgoli, Praveetha Patalay

## Abstract

There are socio-economic inequalities in the experience of mental ill-health. However, less is known about the extent of inequalities by different indicators of socio-economic position (SEP). This is relevant for insights into the mechanisms by which these inequalities arise. For young people’s mental health there is an additional layer of complexity provided by the widespread use of proxy reporters. Using data from the UK Millennium Cohort Study (N=10,969), we investigated the extent to which five SEP indicators (parent education, household income, household wealth, parent occupational status, and relative neighbourhood deprivation) predict adolescent internalising mental health (at ages 14 and 17 years) and how this varies as a function of reporter. Both parent report and adolescent self-report were considered. Regression models demonstrated that whilst greater disadvantage in all five SEP indicators were associated with greater parent-reported adolescent mental health symptoms, only those from more disadvantaged groups of income, wealth, occupational status, and parent education were associated with greater self-reported mental health symptoms. The magnitude of these effects was greater for parent-reported than self-reported adolescent internalising symptoms: SEP indicators jointly predicted 4.73% and 4.06% of the variance in parent-reported symptoms at ages 14 and 17 compared to 0.58% and 0.60% of the variance in self-reported internalising mental health. Household income predicted the most variance in parent reported adolescent internalising symptoms (2.95% variance at age 14 & 2.64% at age 17) and wealth the most for self-reported internalising symptoms (0.42% variance at age 14 & 0.36% at age 17). Interestingly, the gradient and variance explained of parent-reported adolescent mental health across SEP indicators mirrors that of parent’s own mental health (for example, income explained 4.89% variance at the age 14 sweep). Our findings highlight that the relevance of different SEP indicators to adolescent internalising mental health differs between parent and adolescent reports. Therefore, it is important to consider the various perspectives of mental health inequalities gained from different types of reporters.

**Highlights:**

- Income is the most predictive SEP indicator for adolescent internalising mental health (MH).
- SEP indicators explain 0.6% of self and 4.7% of parent reported adolescent MH at 14.
- Socio-economic inequalities in adolescent MH vary by reporter.
- All five SEP indicators are associated with parent-reported adolescent MH.
- SEP indicators have weaker associations with self-reported adolescent MH.

## 1. Introduction

Poor mental health is a leading cause of lost life years (1). It can negatively impact an individual’s life in a plethora of ways (2). Around one in six adults in England present with a common mental health disorder at any one time (3), resulting in an estimated annual cost of £117 billion to the UK economy in 2019 (4). Population-based studies have repeatedly shown socio-economic inequalities in mental ill-health, with individuals lower in socioeconomic position (SEP) faring significantly worse than their more advantaged counterparts (1,5–8). This association has been observed from as young as four years old (9,10).

Adolescence is key when studying how mental health conditions develop and how inequalities manifest. Notably, half of all lifelong cases of mental health problems are present by the age of 14 (11). These difficulties cause long-term economic hardship by reducing school completion rates and hindering the acquisition of social and occupational skills, and are associated with many later life negative outcomes including emotional and marital problems (12). Poor mental health also presents a co-morbidity with physical health issues (13). Consequently, understanding the susceptibility to mental health difficulties conferred by SEP indicators is vital to inform prevention, and to target public health efforts to minimise future adverse impacts. SEP indicators may have different health gradients based on the mental health condition being researched (14). In our study, we focus on internalising difficulties (symptoms of anxiety and depression), as they are the leading cause of disease burden in most high-income countries (15).

### 1.1 SEP indicators and mental health

SEP is a multifaceted construct. Therefore, multiple indicators of SEP are used to measure socio-economic inequalities. These include parent education, household income, household wealth, parent occupational status, and relative neighbourhood deprivation. These indicators are related but cannot be used interchangeably as they confer different resources or difficulties upon individuals (16). Parents with a high level of education have been proposed to recognise their child’s needs more easily and, therefore, know when and how to access structural resources such as mental health treatment (6,17). Household income precipitates access to material resources like healthcare, food, physical environment and housing (18). Therefore, stress, exposure to dangerous situations and food insecurity, among other things, are income dependent. Household wealth, which is only weakly correlated to income (19) and presents much greater inequalities (20), captures both current possessions and past accumulation (21). Wealth reflects parents’ ability to choose environments most beneficial to their children’s long-term success. Therefore, wealth may act on children’s emotional difficulties in a cumulative, rather than an immediate, manner (22). Parent occupational status has a role in positioning individuals (23), and thus their families, in the social structure, hence affecting adolescent exposure to risks and resources (24). Finally, indices of multiple deprivation (IMD) are an official measure of relative neighbourhood deprivation in the United Kingdom (25). Lower perceived safety and reduced levels of neighbourhood cohesion and support in more deprived neighbourhoods may increase an individual’s susceptibility to mental health difficulties (26). The impact of SEP on the onset, severity and course of mental health difficulties is likely to vary by the SEP indicator used (6,9). Therefore, choosing the most relevant indicator for research and policy is vitally important (24). Despite the theoretical differences between these five SEP indicators, very little empirical investigation into their potentially independent impacts on mental health has been made to date.

### 1.2 Multiple reporters of adolescent mental health

There is a concerning lack of consistency in assessments of adolescent internalising mental health between different reporters. In population-based data, these reports typically come from one of four sources: adolescent self-report, teacher report, parent report or clinical interviews. Previous research has shown a weak correlation (0.27) between child and parent reported symptoms (27), with adult reporters consistently underreporting adolescent internalising problems (28). This may be because parents and teachers are not aware of the presence of internalising symptoms unless the adolescent chooses to disclose them (28). Assessment may also vary based on the SEP of the reporter and reportee. Indeed, systematic differences in parent, teacher and self-reports of adolescent mental health have been reported, with greater income-health gradients recorded from adult reports than self-reports (29,30). These disparities bring into question the inferences made in mental health research when only one reporter of mental health is considered. There has been no work to date looking at the relative impact of different SEP indicators on internalising mental health, and whether changes occur based on the reporter. We will focus on the difference between parent and self-reports, as disagreements tends to be higher between adolescents and adults and these are the most commonly used assessors in adolescent mental health research (30).

### 1.3 Objectives

Using a large, nationally representative UK birth cohort, we aim to meet two primary objectives and one secondary objective. The first objective is to assess whether each indicator of SEP (parent education, household income, household wealth, parent occupational status, and relative neighbourhood deprivation) is uniquely associated with internalising mental health in adolescents in the UK. Our second objective is to evaluate whether the estimated magnitude and significance of the association between SEP indicators and adolescent internalizing mental health varies by the reporter (parent or adolescent). To help further unpack any discrepancies seen by reporters we examine whether observed SEP inequalities in adolescent mental health are similar for their parent’s own mental health. We will also examine whether disparities between reporters are present when cohort members were aged 17, in addition to our focus on adolescents at age 14. In the age 17 data, the same measurement is used for both parent and adolescent reported internalising symptoms, this allows us to appraise the role of the survey measure used in our findings.

## 2. Methods

### 2.1 Participants

Data are from the Millennium Cohort Study (MCS), a longitudinal national birth cohort in the United Kingdom (31). The MCS includes data from 19,244 individuals born between September 2000 and January 2002. For further information about this cohort study, see https://cls.ucl.ac.uk/cls-studies/millennium-cohort-study/. This cohort contains detailed information on five different SEP indicators: parent education, household income, household wealth, parent occupational status, and relative neighbourhood deprivation. This information was taken from the age 11 sweep, which is the earliest sweep to include all five SEP variables and precedes the age 14 and age 17 sweeps when the mental health outcomes included in this study were assessed. The age 14 sweep is the first to contain both parent and adolescent reported adolescent internalising mental health. The age 17 sweep is the first to utilise the same measurement for both parent and adolescent reported adolescent internalising mental health.

Our analytical sample considers all cohort members with a at least one measure of SEP recorded at age 11 and at least one measure of mental health recorded at age 14 (N=10,969). In a small number of families where there are more than one cohort members due to multiple births, one member has been selected at random for inclusion in our sample to prevent bias due to the within household nesting effect (32).

### 2.2 Variables

#### Outcomes: Internalising mental health

##### Parent Adolescent Report

Parents reported on their teenager’s mental health using the Strength and Difficulties Questionnaire (SDQ) in both the age 14 and age 17 sweeps (33). The SDQ is a 25-item instrument used to assess emotional, social, and behavioural functioning; it is a widely used research tool for young person mental health and consists of five subscales. In our analysis, we will focus exclusively on the score for emotional symptoms. Higher scores indicate greater mental health difficulties. The scoring is split into a 4-band categorisation: 0-3 represents close to average, 4 slightly raised, 5 to 6 high and 7-10 very high (34). The parent report version of this instrument is a valid screening tool for psychosocial problems (35).

##### Adolescent Self Report

Adolescents reported on their mental health using the Short Moods and Feelings Questionnaire (SMFQ) in the age 14 sweep (36). The SMFQ is a 13-item instrument used to assess adolescent depressive symptoms, a key reflection on internalising mental health difficulties. Answers to the questions are summed to generate a score ranging between 0 and 26. The adolescent self-report version has been shown to have strong internal consistency and be a valid screening tool (37). Higher scores are associated with greater depressive symptoms. Although there is no prescribed cut-off, the optimal value for differentiating depressed and non-depressed cases has been reported as ≥12 (37). In the age 17 sweep, adolescents reported on their mental health using the self-report version of the Strengths and Difficulties Questionnaire, a first-person adaption of the measure used by parents. The 4 band categorisation is slightly different to that of the measure used by parents: 0-4 represents close to average, 5 slightly raised, 6 high and 7-10 very high (34).

##### Parent Self Report

Parents reported on their own mental health using the Kessler Psychological Distress Scale in the age 14 sweep (38). The Kessler scale is a 10-item questionnaire, scored from 0 to 50. It is used to assess anxiety and depressive symptoms experienced in the past four weeks. In this study we used measures completed by the same parent who completed the parent-reported assessment of the adolescent.

#### SEP indicators

SEP indicators, excluding relative neighbourhood deprivation which is a government statistic harvested from one’s postal address, were reported by parents.

National Vocational Qualification (NVQ) levels, which encompass both academic and vocational qualifications, were used to indicate parent education and coded from one to six (1 refers to no qualifications, 2 to GCSE below grade C equivalent, 3 to GCSE grade A-C equivalent, 4 to AS and A level equivalent, 5 to First Degree and 6 to Higher Degree equivalent). We considered the highest parent education achieved for the main responding parent, which in over 90% of cases is the mother.

Parent reports of joint net household income within 20 categories were utilised to indicate household income, these were converted into quintiles for analyses.

Household wealth was derived from four variables: Outstanding mortgages were subtracted from house value to give a measure of *housing wealth*. Debts owed were taken from the amount of investments and assets, to give a measure of *financial wealth*. Housing wealth and financial wealth were then summed to give an overall measure of *total net wealth*. For those with missing responses to one or more of these variables, and where individual’s responses to other questions in the MCS indicate as such (for example, where housing tenure variables indicated no home ownership), a value of zero was given. This approach was based on one that has been used elsewhere with this cohort (22). The household wealth variable was then split into five quintiles for analyses.

The highest parent occupational status was considered. This was operationalised using the National Statistics Socio-economic Classification (NS-SEC) 5 categories (plus a category for those not in employment): managerial, administrative, and professional occupations; intermediate occupations; small employers and own account workers; lower supervisory and technical occupations; semi-routine and routine occupations; and unemployed.

Local relative neighbourhood deprivation was measured from Indices of Multiple Deprivation (IMD) deciles. IMD deciles range from the most deprived decile to the least deprived decile. IMD are an official measure produced for small areas in the United Kingdom known as Lower-layer Super Output Areas (25).

### 2.3 Data analyses

Descriptive statistics of SEP indicators, mental health scores, and mental health scores by each SEP indicator were estimated. Differences between the full cohort and our analytical sample were reported. To visualise differences across the SEP distribution, histograms of mental health outcomes were produced for the most and least disadvantaged groups of each SEP indicator. To examine associations between the SEP indicators, we produced a correlation matrix and tested for collinearity.

Missing data in all analyses was accounted for with multiple imputation using chained equations (39). Missing values varied from 0.05% for relative neighbourhood deprivation to 31.1% for household income, with a total of 15.2% missing cells overall (missing values for SEP variables: parent education 3.6%, household wealth 20.6% (for variables that make up the household wealth variable: house value 10.8%, mortgage value 14.4%, amount of investments and assets 28.3% and debts owed 14.8%) and parent occupational status 2.6%). The number of observations missing for each SEP indicator can be found in the Supplementary file, Section 1, Table S3. A combination of factors including a lack of knowledge regarding household income and wealth and an unwillingness to report these in comparison with educational level and occupation status may explain the greater values of missing data for these variables. The data was imputed 25 times, using a range of auxiliary variables (parents age, housing tenure, parents self-reported health, adolescents self-reported health, parents ethnicity, number of parents/carers and parent reported adolescent mental health from the age 11 sweep). We have adopted the Missing at Random (MAR) approach to deal with missing values in our multiple imputation. This assumes that the expansive information we have about other SEP indicators predict the majority of the variation in missing data (40). We applied sampling and attrition weights in our analyses to account for the stratified clustered design, oversampling of certain groups, and missing data due to attrition.

Outcome variables (parent and self-reported adolescent mental health) were standardised (converted to z-scores) before regression analyses were undertaken to allow direct comparison of effect sizes between reporters. The least disadvantaged socioeconomic group for each indicator acted as the reference group. Analyses were undertaken using both the age 14 and age 17 sweeps, unless stated. All analyses were undertaken in Stata 16 (41).

To address our first objective, linear regression models were conducted with each SEP indicator in turn, to measure the unadjusted relationship between each SEP indicator and adolescent mental health at ages 14 and 17. Parent and self-reported adolescent mental health scores were considered as separate outcome variables. Following this, an expanded model (multiple regression) with all five SEP indicators included simultaneously was produced, to establish the unique contribution of each indicator at ages 14 and 17. The variables available in our dataset are thought to have occurred after parental SEP was established, therefore it was not appropriate to adjust for potential mediators on the causal pathway. The significance of the unique contributions, for the age 14 sweep, was tested with a drop-one analysis: a model with all five SEP indicators compared to models with each indicator removed in turn, using log-likelihood ratio tests. Comparing models in this manner permits a test of their ability to explain the underlying data, since it compares the likelihood of two models, given the observed data. If a model with all five SEP indicators is a better fit to the data compared to a model with any one indicator removed in turn, then the SEP indicator that was dropped can be said to account for significant unique variance in the outcome (42), we reported the percent of imputed data sets that each SEP indicator added a unique contribution to. We chose not to use model selection criteria such as Akaike Information Criteria because our models were nested, with the parameters in the smaller model being a subset of the parameters in the five-indicator model, therefore log-likelihood ratio tests were the most applicable means of assessing the unique contribution of any one SEP indicator. Lastly, we compared the R^2^ values of the unadjusted and expanded models at age 14 and 17 to investigate which dimensions of SEP explain more variance than others.

For our second objective, to evaluate whether the estimated magnitude and significance of SEP indicators on adolescent internalising mental health varies by the assessor, the model predicted means of both parent and self-reported adolescent mental health were plotted together for each of our five SEP indicators and their socioeconomic groupings for both ages 14 and 17.

To investigate whether observed SEP inequalities in adolescent mental health are similar for parent’s own mental health, unadjusted linear regression models and model predicted means were applied to parent’s standardised mental health scores (Kessler scale), with each SEP indicator in turn. The predicted means were then plotted with parent-rated adolescent mental health, from the age 14 sweep, for each SEP indicator. The insight this provides will help in understanding the results from our second objective.

In addition, we conducted two further analyses. Firstly, to report the associations of SEP with probable mental health disorders, dichotomised mental health outcomes at age 14 (clinical levels of mental ill-health or not) were produced for both parent report and adolescent self-report measures based on established cut-offs for the respective measures (scores ≥7 on the SDQ (very high (34)) and scores ≥12 on the SMFQ (optimal value for establishing depressed and non-depressed cases (37)). Logistic regressions and plots were used to visualise the unadjusted relationship between each SEP indicator and very high levels of adolescent internalising symptoms. The absence of clinical levels of internalising symptoms was the reference category. Secondly, to determine whether the effect of each SEP indicator was different in males and females as previously reported (43), we conducted a sex-SEP indicator interaction for parent and self-reported adolescent internalising symptoms at age 14. Model predicted means were estimated and plotted for adolescent internalising mental health for both sexes and for each SEP indicator.

## 3. Results

### 3.1 Descriptive Statistics

Differences between the full cohort sample and the selected analytical sample were negligible for most variables (see Supplementary file, Section 1, Table S1). There were no issues of collinearity between the five SEP indicators (see Supplementary file, Section 1, Table S3 and Table S4). A noticeable difference in mean parent-reported adolescent internalising symptoms between the most and least disadvantaged groups, regardless of the age of adolescents or SEP indicator used, was observed. However, this difference was considerably smaller and less consistent across SEP indicators for adolescent reports of internalising symptoms (see Table 1).

**Table 1:**
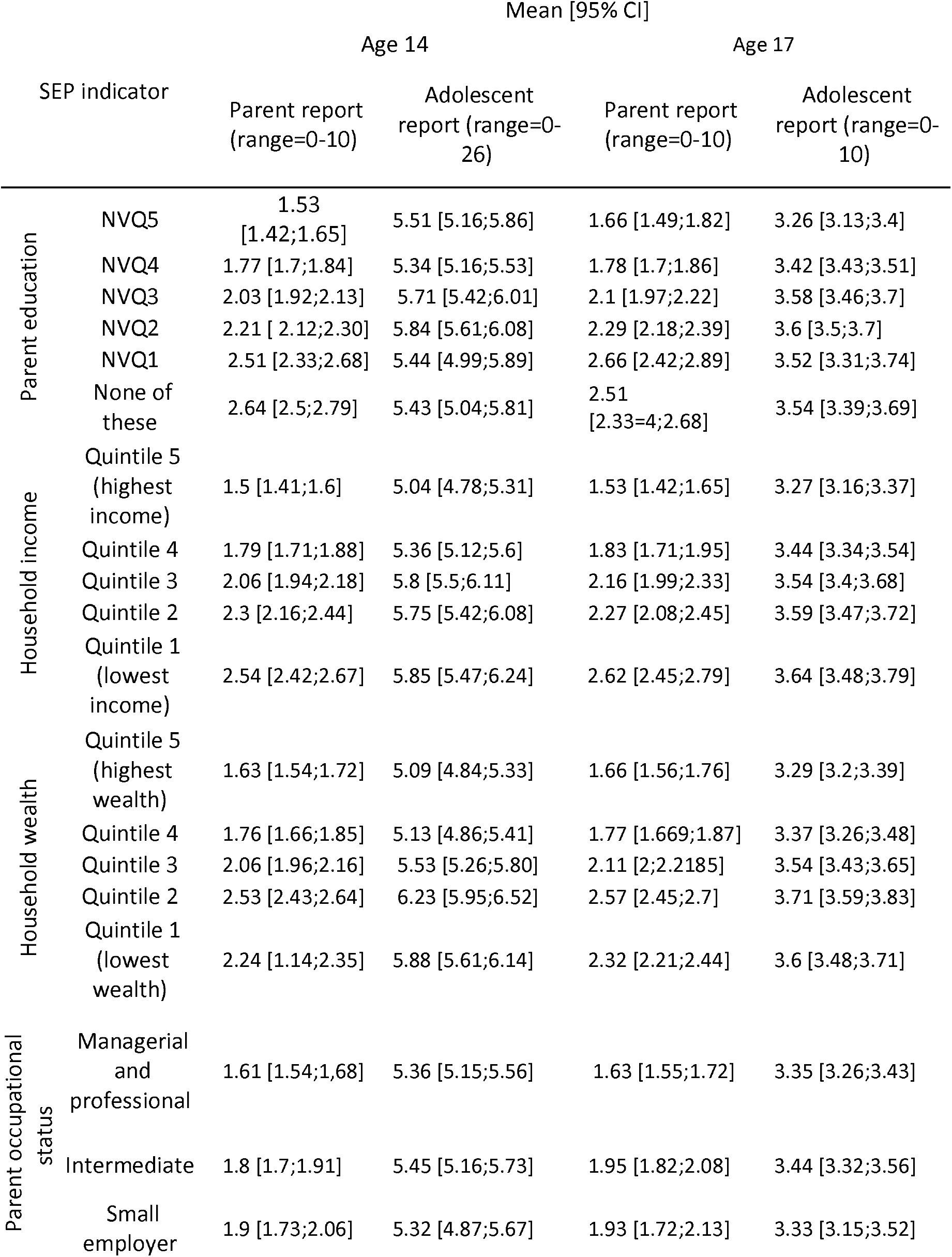
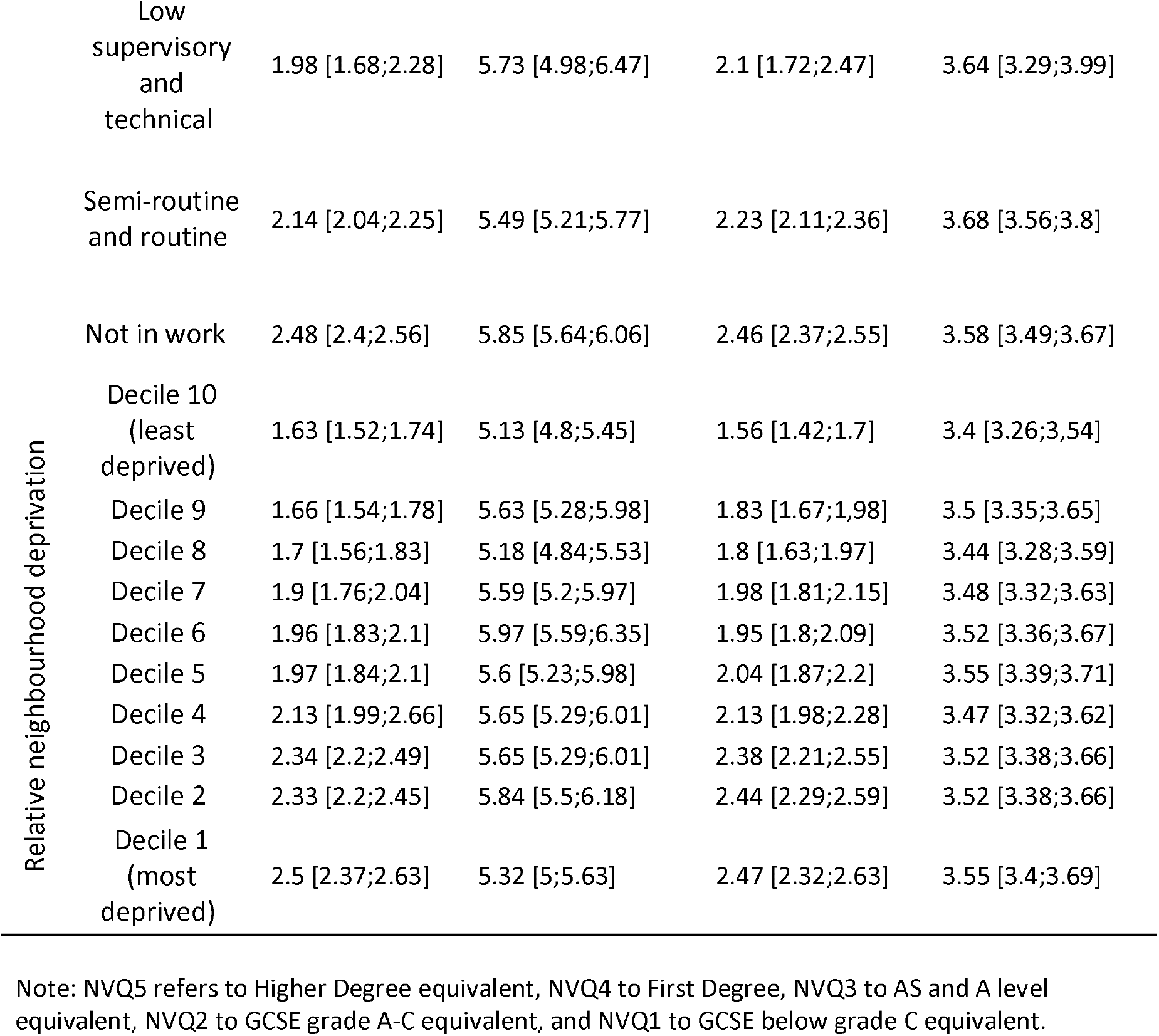
Mean (95% confidence intervals), for parent and self-rated adolescent mental health outcomes at age 14 and age 17 by SEP indicators.

Differences in the distributions of adolescent internalising mental health score (both parent and self-report) at age 14, between the most and least disadvantaged groups for each SEP indicator, can be found in Figures 1a and 1b. The most and least disadvantaged groups have a more similar distribution of internalising symptoms when reported by adolescents, than parents.

**Figure 1a.**
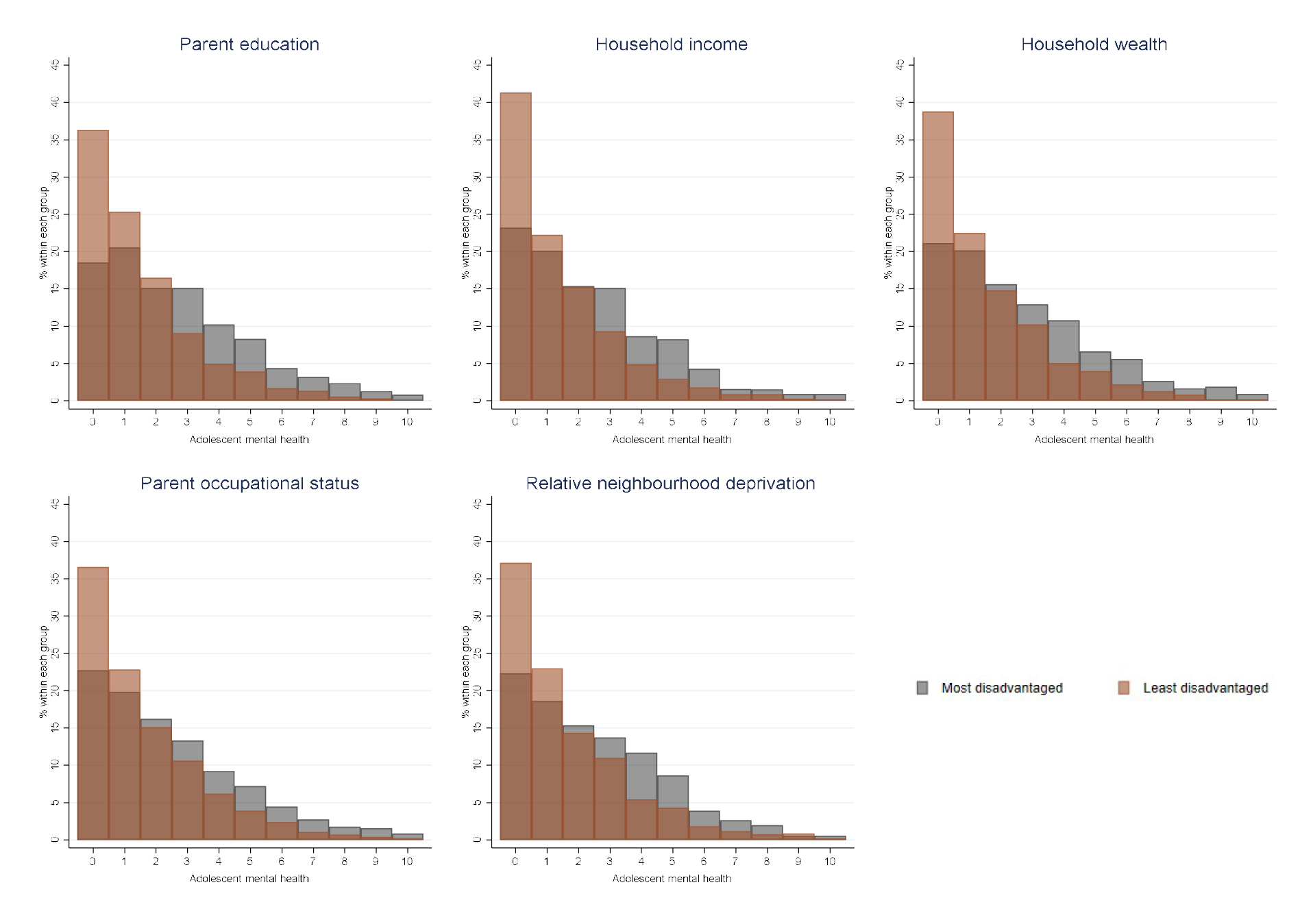
Distribution of parent reported adolescent internalising mental health at age 14, by SEP indicator.

**Figure 1b.**
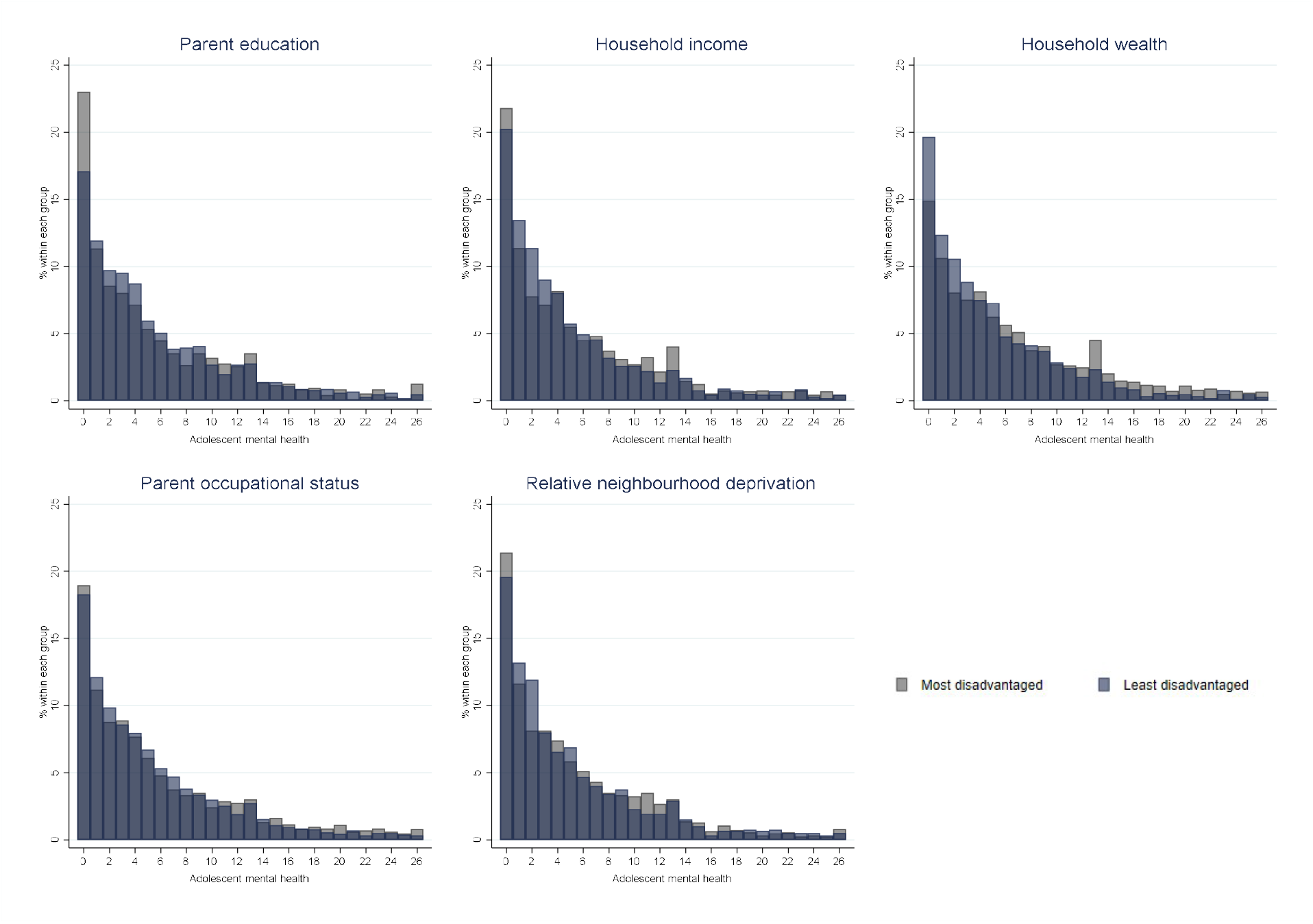
Distribution of self-reported adolescent internalising mental health at age 14, by SEP indicator.

### 3.2 Objective 1. The unique association of SEP indicators with internalising mental health in adolescents

#### Parent reported adolescent mental health

For all SEP indicators considered, we observed an association between being in the most disadvantaged socio-economic group and greater parent reported adolescent internalising symptoms in the unadjusted models. For example, belonging to the lowest household income group was associated with an increase of around half of a standard deviation (b=.49[0.41;0.57]) in internalising symptoms at age 14, compared to those from the highest income group (see Table 2). In the expanded model, an association between the most disadvantaged groups of all SEP indicators considered, excluding relative neighbourhood deprivation and parent education for mental health recorded at ages 14 and 17 respectively, and greater parent-reported adolescent internalising symptoms was detected. Household income presented the largest association with the outcome (at age 14 b=0.2[0.1;0.31] & age 17 b=0.22[0.1;0.35]) (see Table 3). Drop one analyses revealed that all SEP indicators added a unique contribution to the parent-reported model at age 14 after accounting for other SEP indicators (see Supplementary file, Section 2, Table S5).

**Table 2:**
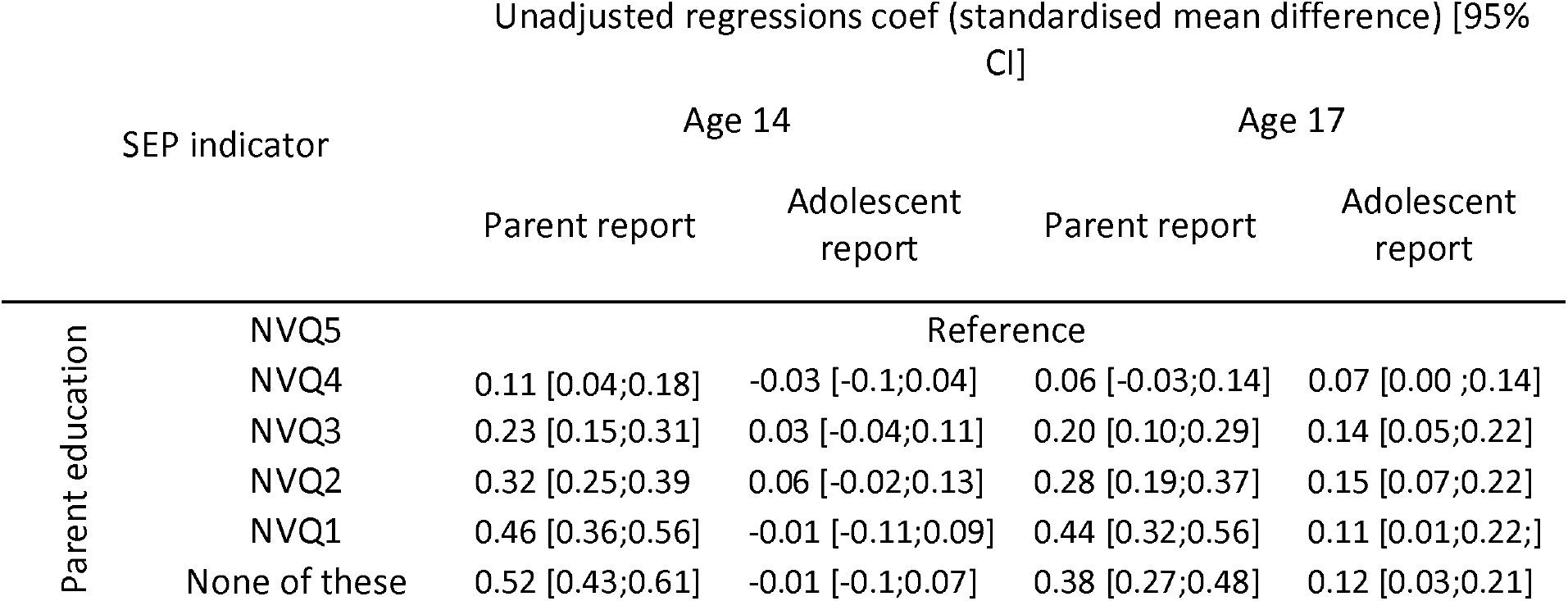
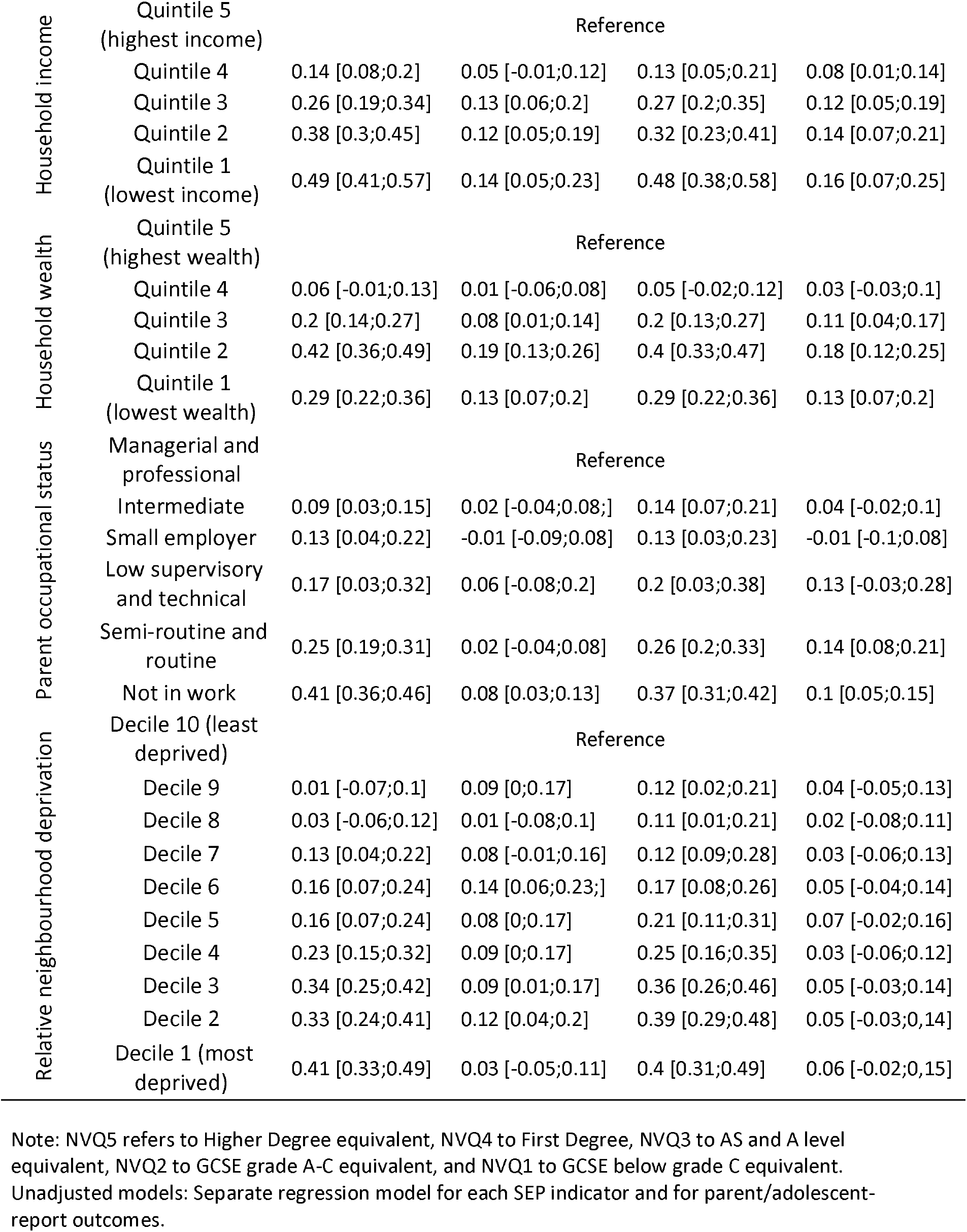
Unadjusted regression coefficients (and 95% confidence intervals) for SEP indicators and both parent and self-reported adolescent mental health ratings

**Table 3:**
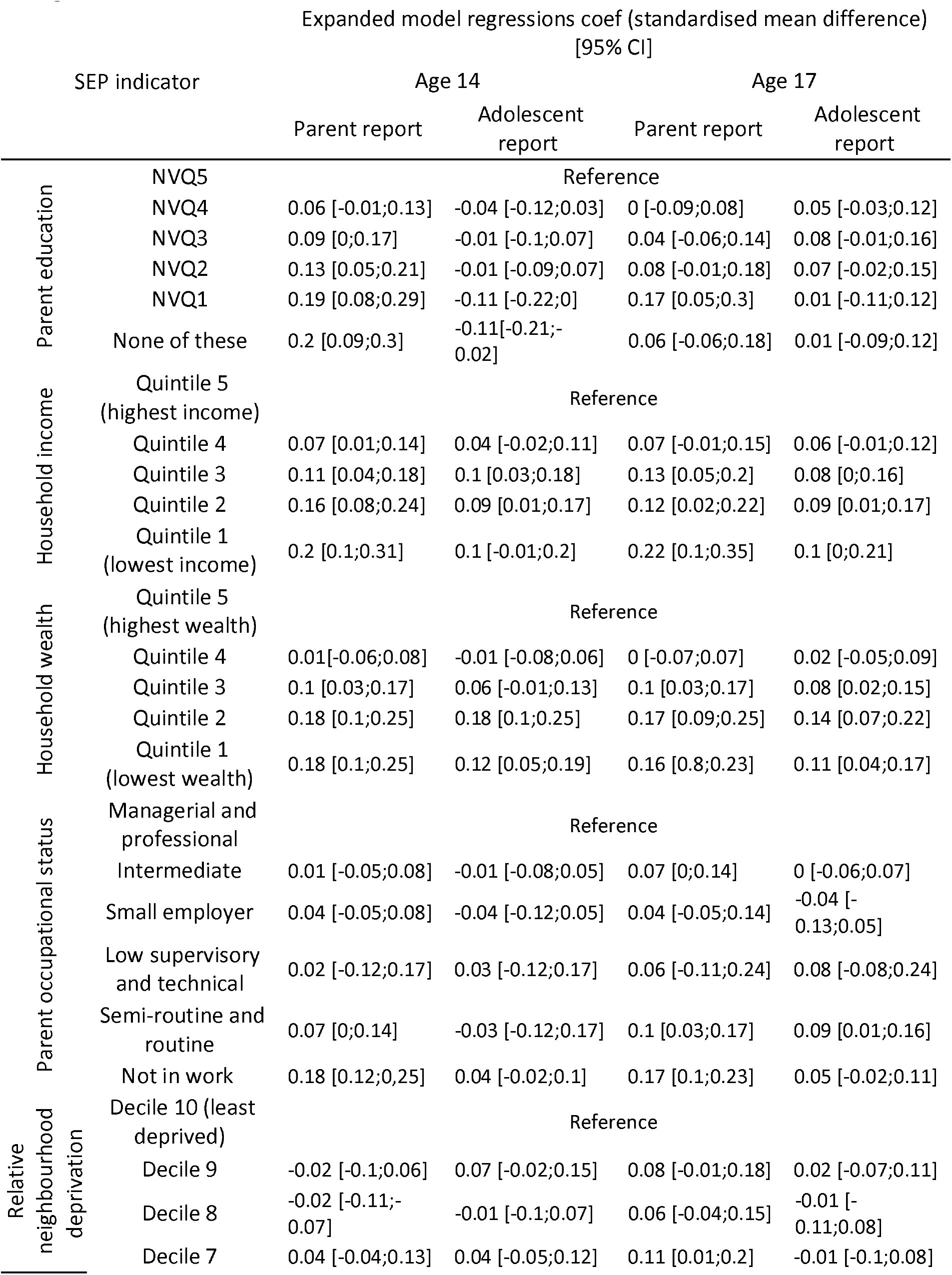
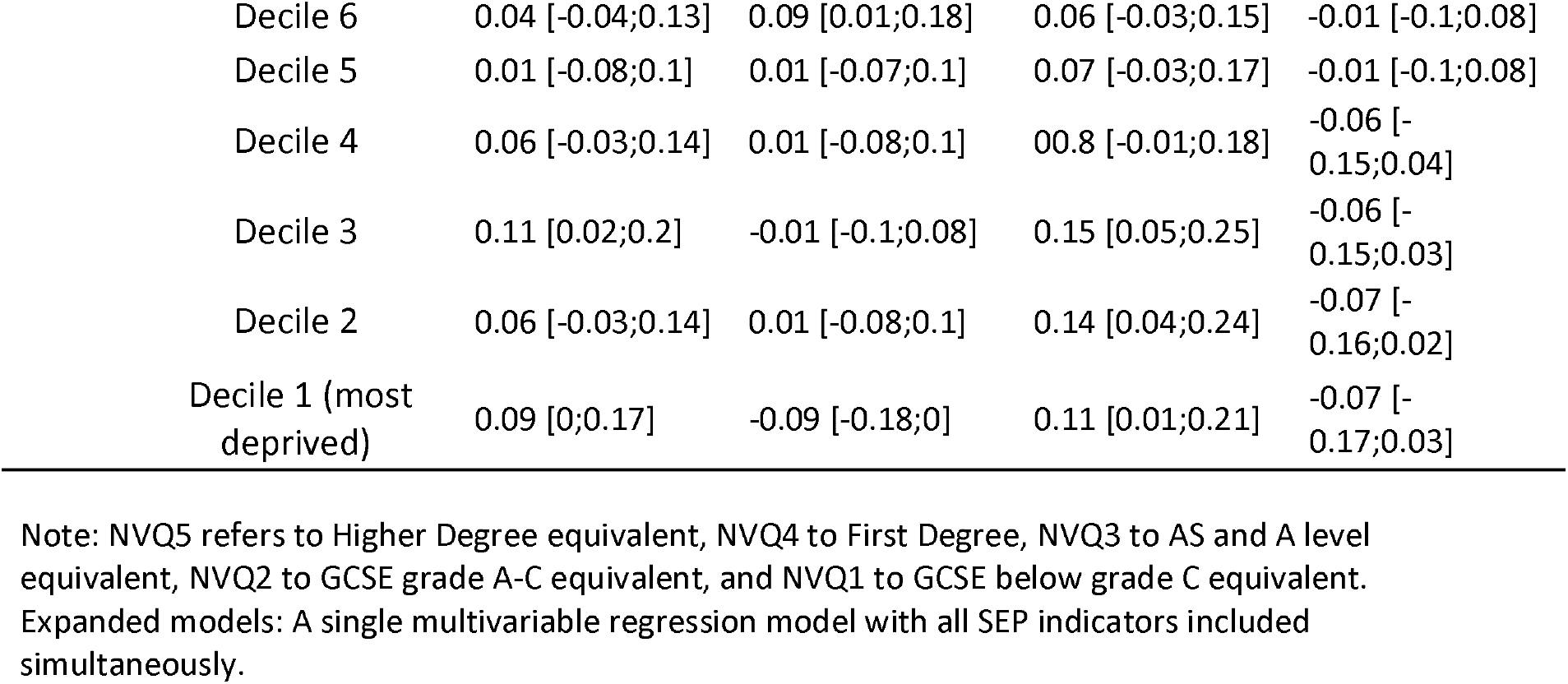
Expanded model (adjusting for other SEP measures) regression coefficients (and 95% confidence intervals) for SEP indicators and both parent and self-reported adolescent mental health ratings

#### Adolescent self-report

In the unadjusted models, those in the most disadvantaged socioeconomic groups of household income (at age 14 b=0.14[0.05;0.19] & age 17 b=0.16[0.07;0.25]), household wealth (at age 14 b=0.13[0.07;0.2] & 17 b=0.18[0.1;0.25), parent occupational status (at age 14 b=0.08[0.03;0.13] & age 17 b=0.1[0.05;0.15]), and parent education (only for mental health recorded at age 17 b=0.12[0.03;0.21]) had greater adolescent self-reported internalising symptoms than those from the least disadvantaged groups. Relative neighbourhood deprivation did not present a consistent association with adolescent self-reported symptoms, whilst parent education was not associated with the outcome at age 14. However, in the expanded model an association was observed only with reduced household wealth and greater self-reported internalising symptoms at ages 14 and 17. Nevertheless, the SEP indicators we have used are correlated (see Table S3), and it is possible that these variables are on the same causal pathway leading to adolescent mental health. Therefore, in our expanded models some of the indicators of SEP may mediate other SEP indicators relationships with adolescent mental health. All SEP indicators added a unique contribution to the adolescent-reported model at age 14 after accounting for other SEP indicators.

Overall, associations between lower SEP and more adolescent internalising symptoms were greater and more consistent for parent-reports than adolescent self-reports. Indeed, SEP indicators explained more variance in the model predicting parent-reported internalising symptoms than in the model predicting self-reported symptoms (see Figure 2a and 2b). We also observed that a model containing all five SEP indicators explained more variance than any single SEP indicator. For both age groups, household income explained the most variance from parent reports, and household wealth the most variance from self-reports. For adolescent mental health at age 14, household wealth explained the least variance from parent reports and parent education the least from self-reports, while relative neighbourhood deprivation explained the least variation from both reports at age 17.

**Figure 2a.**
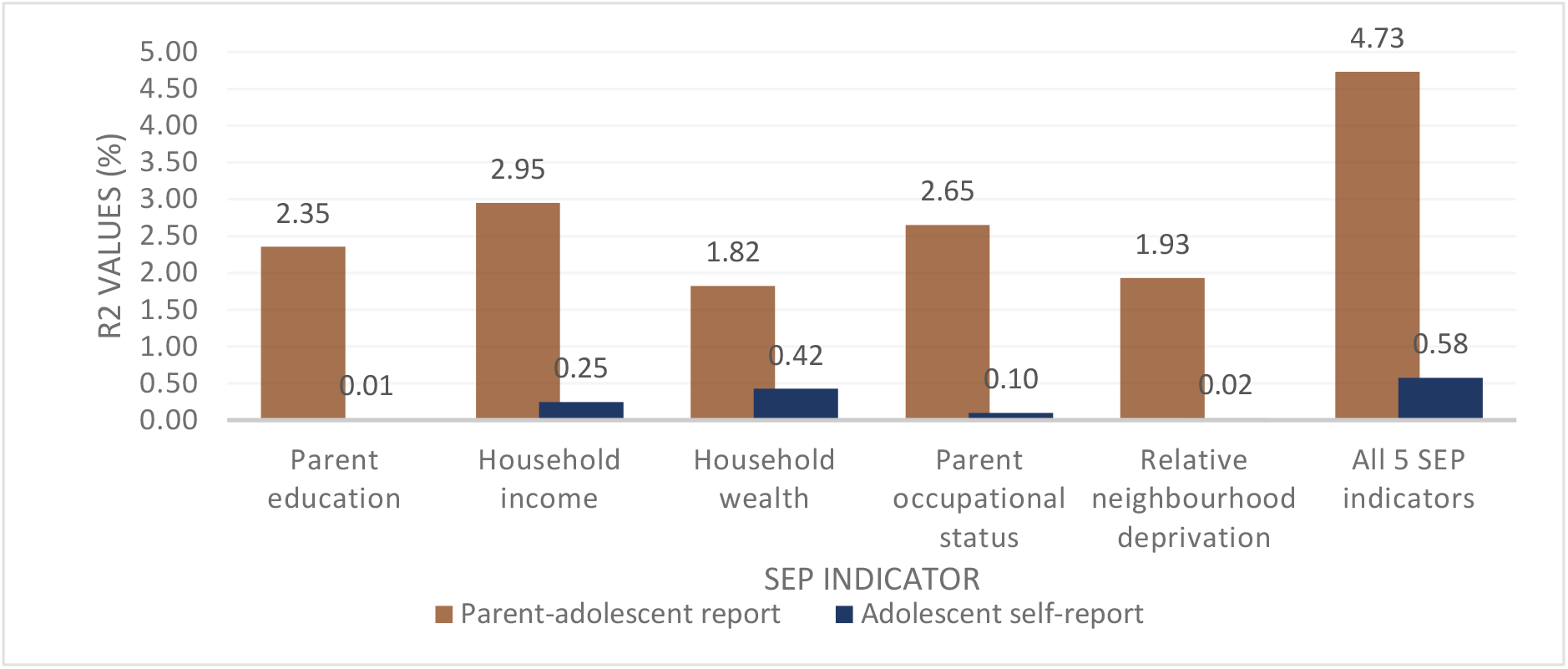
The variance of parent and self-reported internalising adolescent mental health explained by different SEP indicators (R) at age 14.

**Figure 2b.**
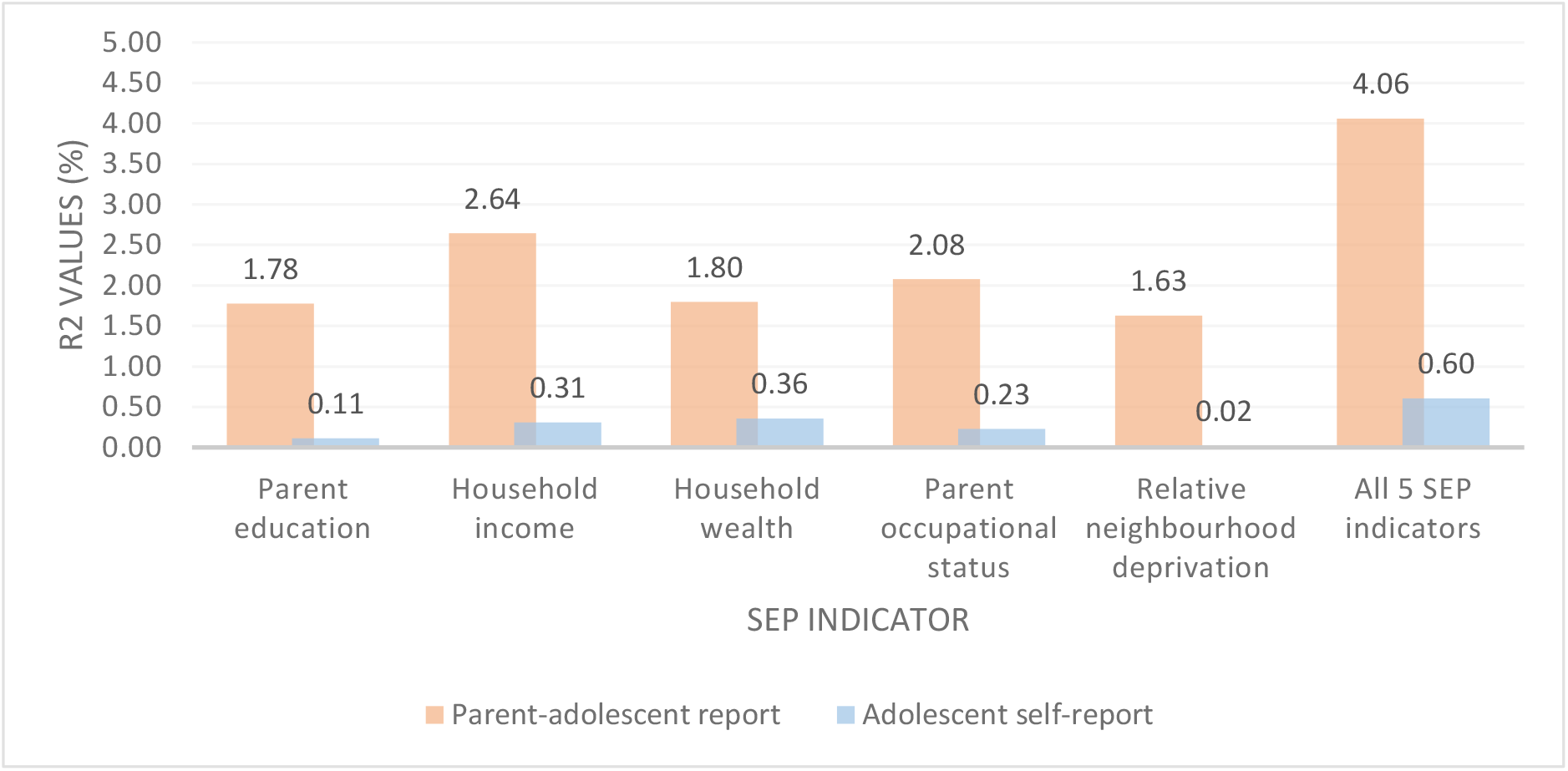
The variance of parent and self-reported internalising adolescent mental health explained by different SEP indicators (R) at age 17.

### 3.3 Objective 2. Comparison of SEP inequalities in adolescent mental health by assessor

For all five SEP indicators considered, we observed that parents from more disadvantaged backgrounds rate their children as having worse mental health than their peers from less disadvantaged backgrounds (see Figure 3). SEP presents a smaller and less consistent association with self-reported adolescent internalising mental health compared to parent-reported adolescent symptoms. Both age groups present very similar patterns for parent adolescent reports and adolescent self-reports on the relationship between SEP indicators and internalising mental health.

**Figure 3.**
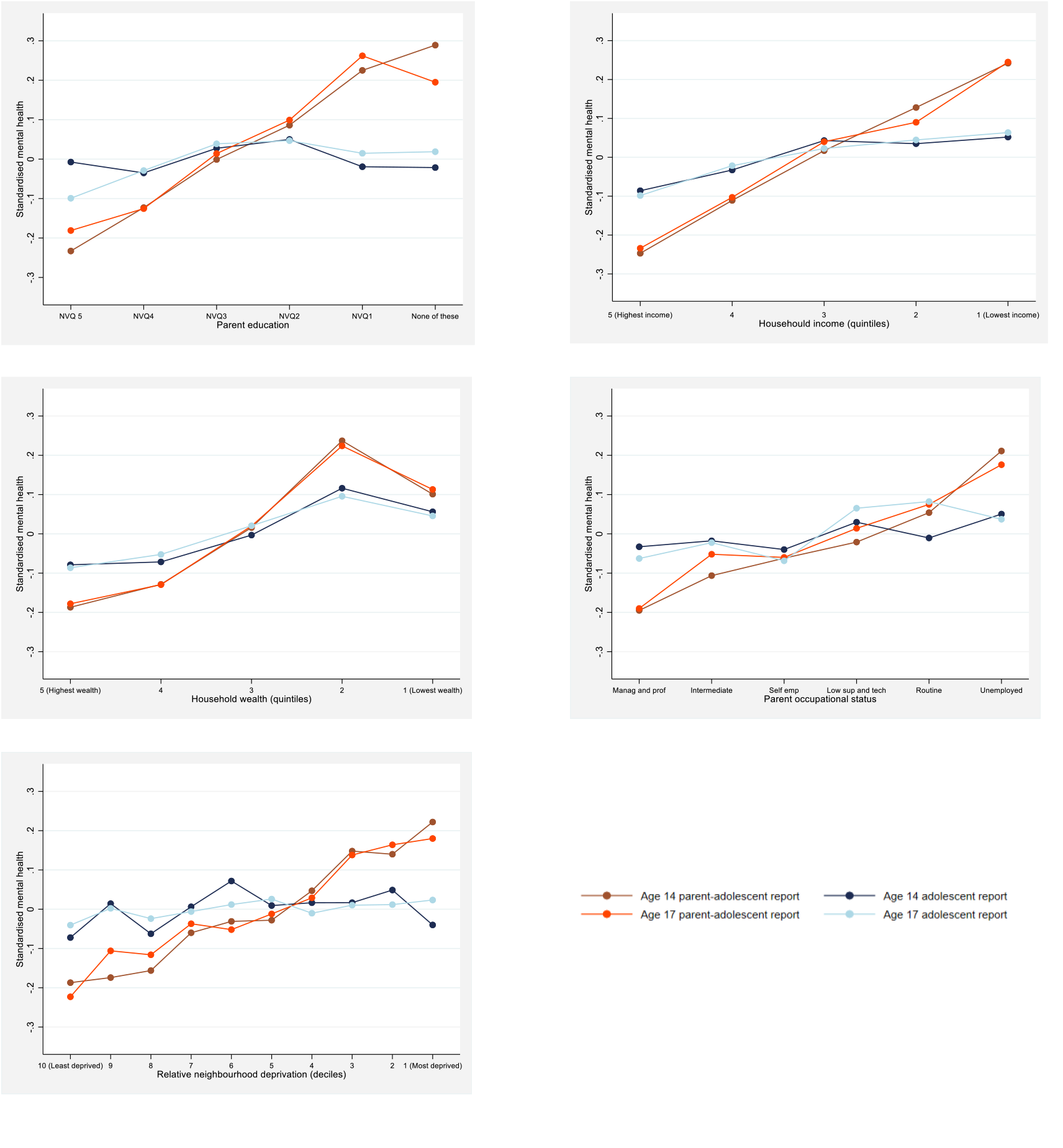
Visualisation of the association between adolescent internalising mental health, SEP indicators and the effects of the reporter.

### 3.4 Comparison of SEP inequalities between parent-reports of adolescent mental health and parents own mental health

There is great similarity between observed SEP inequalities for parent reports of adolescent internalising symptoms, and parent self-reported symptoms (see Figure 4). Parents in the most disadvantaged groups rate their own and their children’s mental health as worse than their peers in less disadvantaged groups, for all 5 SEP indicators. SEP indicators explained more variance in the model predicting parent self-reported internalising symptoms than in the model predicting parent-reported adolescent symptoms (see Supplementary file, Section 3, Figure S1).

**Figure 4.**
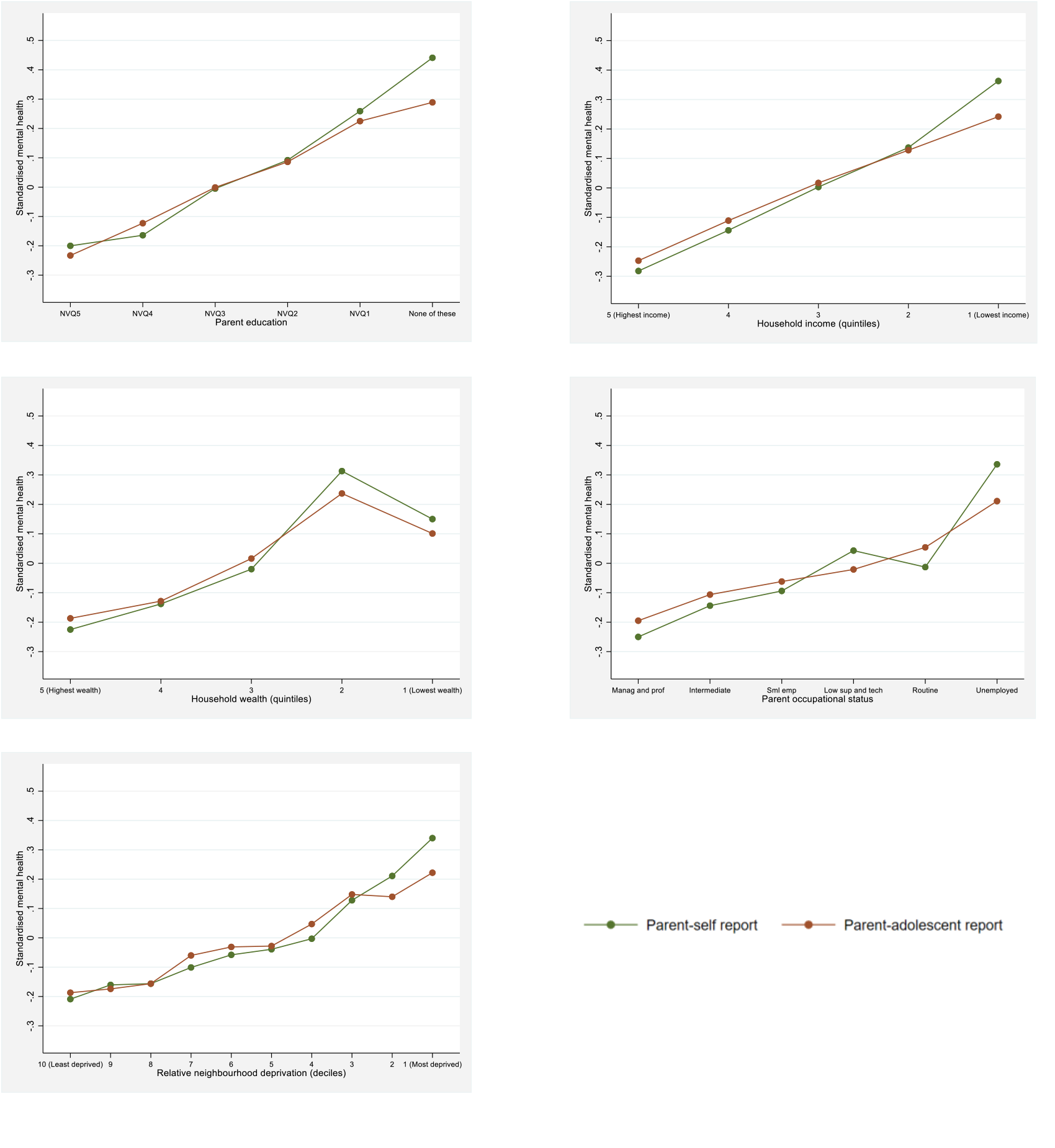
Visualisation of the association between parent self-reports of internalising mental health and parent-reports of adolescent internalising mental health, and SEP indicators.

### 3.5 Planned additional analyses

Tables and figures for these planned analyses can be found in the Supplementary file (Section 4).

#### 1. Adolescent internalising mental health as a binary outcome

Parents reported 462 (4.2%) members of our analytical sample with clinical-level internalising symptoms, whilst self-reports revealed 1,696 (15.5%) individuals with clinical-level symptoms at age 14 based on established cut-off scores for measures used in this study. Models showed patterns similar to the main results (see Supplementary file, Section 4, Figure S2). For all five SEP indicators, an association was observed between lower SEP and increased clinical-levels of parent-reported adolescent internalising symptoms. For parent education, for example, adolescents from the most disadvantaged group had a 249% increase in the odds (OR=3.49[2.11;5.77]) that their parent would report them as having clinical-level internalising symptoms, compared to those from the least disadvantaged group. The relationship with relative neighbourhood deprivation was weakest (OR=2[1.29;3.09]). For adolescent self-reported clinical-level internalising symptoms, an association was seen only for the most disadvantaged groups based on income (OR=1.36[1.08;1.71]), wealth (OR=1.42[1.17;1.73]) and occupational status (OR=1.31[1.14;1.5]).

#### 2. Sex and SEP interaction

We found that females are consistently rated as having more internalising symptoms than males, regardless of the reporter. The difference in mental health scores between the sexes at age 14 was greater when reported by adolescents than by their parents. With regards to the interactions between sex and SEP indicators, we observed similar patterns to the main results, regardless of sex, for the association between SEP and both parent and self-reported adolescent internalising symptoms (see Supplementary file, Section 4, Figure 3a and 3b). In other words, the associations between SEP and mental health were not discernibly different in males and females at this age.

## 4. Discussion

As growing evidence points to the long-term effects of mental ill-health in adolescence on outcomes such as adult health, academic achievement, and human capital accumulation (44), identifying the socioeconomic antecedents of adolescent mental health is an essential objective of policy and research. In this paper, we have used a large, nationally representative UK birth cohort to investigate the extent to which five objective SEP indicators (parent education, household income, household wealth, parent occupational status, and relative neighbourhood deprivation) predict adolescent internalising mental health and how this varies as a function of the reporter.

All five SEP indicators were associated with parent-reported adolescent internalising symptoms, while only household income, household wealth, and parent occupational status were found to be associated with adolescent self-reported mental health at both ages 14 and 17. Household income was the most important SEP indicator for parent reported mental health, while household wealth alongside household income were the most important for self-reported mental health. Our findings also highlight that the inferences made about the health gradient in internalising adolescent mental health depend on who assesses the adolescent’s mental health. The systematic differences we observed between parent and self-reported evaluations suggest that the estimated magnitude and significance of the health gradient was much greater when parents reported adolescent internalising symptoms, compared to when adolescents reported on themselves. Notably, the gradient of parent-reported adolescent internalising symptoms across SEP indicators mirrors that of the gradient for parent ratings of their own mental health.

### 4.1 Different SEP indicators and adolescent mental health

Income is the SEP indicator most utilised in research regarding health gradients (8,29,30,45,46) and a lower income is consistently associated with worse adolescent mental health. Our research has reaffirmed this: of our five SEP indicators, income explained the most variance for adolescent internalising mental health when reported by parents and the second most when self-reported. Parents with a high income are able to access appropriate medical care and modify their child’s environment to reduce the severity of their symptoms, which is likely more difficult for families with a lower income (47). The former is especially notable in the UK, where two-thirds of patients experience a year-long wait for NHS provided mental health services (48). Alternatively, the Family Stress Model (FSM) posits that a reduction in, or low income constitutes a risk for child mental health, through economic pressure (e.g., difficulty paying rent and shopping habits in food and clothing), and negative changes in parental mental health, marital interaction and therefore parental quality (49). For example, a recent review found parents who are stressed due to a low income show harsher parenting, are less supportive, and present a lower provision of social and cognitive enrichments for their child (50). Additionally, a study utilising the MCS demonstrated that transitions to income-poverty in childhood increase the odds of child and maternal mental health problems, independent of changes in employment status (51). The FSM is also purported to be relevant to parents educational differences in SEP as well as economic differences (52).

A strong association was observed between parent education and parent-reported adolescent mental health. Indeed, adolescents whose parents had no formal qualifications were 3.49 times more likely to have disorder-levels of parent-reported internalising symptoms at age 14 (determined by clinical cut offs), than their peers from households with a higher degree. This supports patterns seen in a study in Norway (53), but existing research is inconsistent, with an analysis of an American cohort, suggesting parent education predicts severity, but not onset, of internalising symptoms (54). High parent education is likely to confer better access to resources, such as mental health treatment (6). It may also signify parents’ patience and nurturing qualities, and hence their parenting practises (47). However, we only observed a relationship between parent education and adolescent self-reported mental health in an unadjusted model at age 17 but not at age 14, and in an expanded model, reduced parent education was associated to *fewer* self-reported internalising symptoms at age 14. Parent education, and the advantages it may confer, might be of less importance to adolescents as peer-group influences increase (55) and, thus, may not be incorporated into adolescents’ expectations of their own mental health. The lack of a consistent relationship between parent education and adolescent mental health in the expanded model, is likely explained by the inclusion of other SEP indicators (e.g. income) which may be mediators of the association between education and mental health.

With regards to the other SEP indicators, we observed a weak relationship between household wealth and adolescent internalising symptoms when reported by parents but a relatively strong relationship when symptoms were self-reported. A previous study in the US found a stronger relationship between wealth and disorder-level internalising symptoms than suggested by our study (56). However, financial assets (one half of the wealth variable) have previously been found not to be independently associated to this outcome in the UK (22), unlike in the USA. This is most likely explained by the higher frequency of financial asset ownership in the USA compared with the UK (57). Housing wealth may predispose families to their neighbourhoods and schooling conditions, as house prices are greater in the catchment areas of more desirable schools (58).

Compared to the other indicators, parent occupational status was moderately predictive of adolescent internalising symptoms across both parent and self-reports. Recent comparable research is limited. Occupational status likely reflects either material and structural factors (perhaps due to income), or a hierarchy of power and prestige in society (social class). However, with the rise of new jobs that do not directly correspond to NS-SEC occupational categories (e.g. influencer and life coach), and with most measures being developed and validated on men (24), it has been argued that occupational classification systems are quickly becoming obsolete (59). Indeed, only 2% of parents in our analytical sample worked in low supervisory or technical occupations.

Relative neighbourhood deprivation was predictive of adolescent internalising symptoms when reported by parents, but no such relationship was present when self-reported by adolescents. It is unclear whether those living in communities characterised by more disadvantage face greater mental health difficulties due to living in such an environment (social-interactive, environmental, and institutional mechanisms (60)), or because they tend to have reduced financial resources and education (26). Despite this, we found that relative neighbourhood deprivation did add a unique contribution to our models. However, a recent review reported that only 9 of 18 studies (50%) observed an association between neighbourhood deprivation, particularly neighbourhood social environment, and young people’s internalising mental health (26). Another study reported that associations between changes in neighbourhood deprivation and mental health disappeared after controlling for other life events (61).

Our study has shown that although females have greater internalising symptoms, the socioeconomic gradient is in fact consistent between males and females, regardless of the reporter. Previous research related to symptom development between these ages has found no association between internalising symptoms and income in adolescent males, but such an association in adolescent females (45). A disparity in the health gradient between the sexes has also been observed in later life (62).

### 4.2 The difference in SEP gradients between assessors

Differences in the ratings of adolescent internalising mental health between assessors are well-established (27). However, only two studies, to our knowledge, have focused on the adolescent mental health gradient and whether this varies by the assessor, but both utilised only one (income) SEP indicator (29,30). They found that the differences between respondents’ ratings of British and Australian adolescents (11 to 15 and 10 to 15-year-olds, respectively) internalising mental health to vary based on income, with adolescents’ own assessments of their mental health suggesting a lower income-health gradient than their parents’ assessments. The latter study found that the magnitude of difference reduced when maternal mental health variables were controlled for, but these may be on the causal pathway (29). The confirmation of these patterns in our study, for multiple SEP indicators, suggests these findings may be due to a genuine mechanism of systematic bias between assessors in the evaluations of adolescent mental health across SEP, rather than being specific to the data or methods used. Additionally, our inclusion of age 17 data, with parents and adolescents reporting mental health using the same measure, established the consistency of our findings between the age groups. This allows us to confirm that the differences we have observed between reporters at age 14 were unlikely to be driven by slight differences in the mental health measures used at this age.

For use in research and policy, it is important to evaluate the relative utility of each of the assessors’ viewpoints, but this is hard to distinguish in the absence of an ‘objective’ measure. The aforementioned British study attempted to deconstruct the predictive power of assessors using the number of days a child was absent from school (30). They found that only teacher reports of internalising symptoms were associated with this measure. However, absenteeism may not be the most reliable measure of mental health. Absentee rates reported by teachers may be clouded in the same type of socially graded heterogeneity as mental health reports (29). Furthermore, some children’s home environments may exacerbate their symptoms, and so they prefer to attend school. Crucially, it is not acceptable to ignore self-reports of adolescent mental health. Children as young as seven have been shown to be reliable self-reporters of their own health (63) and mental health is inherently subjective, thus an adolescent who feels distress is no less valid in their report just because their parents have not noticed. Indeed, parent reports on their adolescent’s mental health could simply reflect their own mental health states, thus explaining the similarity in SEP inequalities between parent reported adolescent mental health and parent’s own mental health observed in our study. The greater inequalities in parents self-reports than adolescents self-reports of internalising symptoms, could then reflect only the accumulation of adverse effects from living in a more disadvantaged SEP over time (47) and its effect on mental health. However, the consistency between our findings at age 14 and age 17 suggests this may not begin until individuals become adults and their own socio-economic circumstances become more salient compared to their parent’s socioeconomic circumstances.

It is possible that the true extent of the adolescent-reported health gradient has not been captured by our use of objective SEP indicators. A recent study of multiple European countries found that inequalities in self-reported health and life satisfaction were larger when adolescent subjective social status (SSS) rather than objective SEP measures was used (59). SSS reflects relative standings (in school or neighbourhood, for example) rather than absolute levels. The subjective nature of SSS might allow it to share bidirectional effects with health (64). Objective SEP indicators did not account for the association between SSS and mental health, and indeed SSS and objective SEP indicators were reported to share just 6-8% of common variance (59).

Despite the greater prevalence of clinical diagnoses in more objectively disadvantaged groups (6), objective SEP indicators are likely less relevant to adolescents than SSS. Adolescent’s expectations, and thus ratings, of their mental health may be more concerned with local and subjective comparisons, and thus perceptions and psychosocial processes, than societies material inequalities. As individuals age and become more self-dependent, they may incorporate a greater awareness of population inequalities into their expectations, and thus ratings, of their own and their children’s mental health. This could explain why socio-economic inequalities in adolescent mental health, measured with objective SEP indicators, are greater when reported by parents than by adolescents themselves. However, we did observe inequalities in adolescent self-reported mental health for three objective SEP indicators. Perhaps subjective and objective SEP indicators relate to adolescent mental health through different causal pathways (65).

### 4.3 Strengths and limitations

Our study was supported by the strength of a large, nationally representative, longitudinal cohort, which has collected both parent and self-report measures of internalising adolescent mental health and information on five objective SEP indicators. Our inclusion of the age 17 sweep and its utilisation of the same measure for parent adolescent and self-reports of mental health allowed us to confirm that our findings at age 14 are unlikely to be measure driven.

However, our study has several limitations which are inherent in the data source and how the outcomes were measured. Firstly, bias in attrition, where higher rates of attrition have been reported for adolescents of low SEP or those with mental health difficulties, most likely led to an underestimation of the health gradient. Another limitation is the high levels of missing data for some SEP indicators. This may have led to a reduction in the precision of our results, and increased the difficulty of health gradient comparisons across SEP indicators (66). The Missing at Random assumption behind the multiple imputation is based on the rich auxiliary information available in the dataset, and although this method is better than listwise deletion of data (67), the meeting of the MAR assumption cannot be formally confirmed. Additionally, potential confounders of the relationship between SEP and adolescent mental health (e.g. genetics) were not adjusted for in this analysis, as they were not available to us in this dataset. The findings also do not address the possible mechanisms for the health gradient seen, but instead describe the health gradient and observed associations.

### 4.4 Conclusions

This study has found that five objective SEP indicators were associated with parent-reported adolescent internalising mental health in a nationally representative cohort. The mental health of adolescents in more disadvantaged groups was rated worse than their peers from less disadvantaged groups, thus producing a health gradient. Interestingly, the parent’s mental health gradient mirrored that of the parent-reported adolescent mental health gradient. When using adolescent self-reports, only three of the five SEP indicators (household income, household wealth, and parent occupational status) were uniquely associated with internalising symptoms when adjusting for the other indicators. The estimated magnitude and significance of the health gradient was larger when rated by parents than by adolescents themselves for five SEP indicators. Thus, a systematic bias existed in estimates of the adolescent mental health gradient dependent on the assessor. Additionally, household income was the most important SEP indicator for parent reported adolescent mental health, while household income and household wealth were the most important indicators for adolescent self-reported mental health.

## Data Availability

Cohort data comply with ESRC data sharing policies, readers can access data via the UK Data Archive.

https://www.data-archive.ac.uk/

## Acknowledgements

We are grateful to the cohort members of the Millennium Cohort Study and their families. We are also grateful to the Centre for Longitudinal Studies (CLS), for the data collection, and to the UK Data Service for making them available. Neither the CLS or UK Data Service have any responsibility for the analyses or interpretation of this data.

## Author contributions

Conceptualisation: MH,

PP Methodology: MH, ET, HHB, PP

Analysis: MH

Visualisation: MH

Supervision: PP, ET, HHB

Original draft: MH

Review and editing: MH, ET, HHB, PP

Revision: MH, ET, PP

## References

1. Murray CJL, Vos T, Lozano R, Naghavi M, Flaxman AD, Michaud C, et al. Disability-adjusted life years (DALYs) for 291 diseases and injuries in 21 regions, 1990-2010: A systematic analysis for the Global Burden of Disease Study 2010. Lancet. 2012;

2. Copeland WE, Wolke D, Shanahan L, Costello J. Adult functional outcomes of common childhood psychiatric problems a prospective, longitudinal study. JAMA Psychiatry. 2015;

3. Stansfeld S, Clark C, Bebbington P, King M, Jenkins R, Hinchliffe S. Mental health and wellbeing in England: Adult Psychiatric Morbidity Survey 2014. In: Adult Psychiatric Morbidity Survey 2014. 2016.

4. McDaid D, Park A., Davidson G, John A, Knifton L, McDaid S, et al. The economic case for investing in the prevention of mental health conditions in the UK [Internet]. 2022. Available from: https://www.mentalhealth.org.uk/sites/default/files/2022-06/MHF-Investing-in-Prevention-Report-Summary.pdf

5. Marmot M, Bell R. Fair society, healthy lives. Public Health. 2012;

6. Reiss F. Socioeconomic inequalities and mental health problems in children and adolescents: A systematic review. Social Science and Medicine. 2013.

7. Meyer OL, Castro-Schilo L, Aguilar-Gaxiola S. Determinants of mental health and self-rated health: A model of socioeconomic status, neighborhood safety, and physical activity. Am J Public Health. 2014;

8. Jacquet E, Robert S, Chauvin P, Menvielle G, Melchior M, Ibanez G. Social inequalities in health and mental health in France. The results of a 2010 population-based survey in Paris Metropolitan Area. PLoS One. 2018;

9. Davis E, Sawyer MG, Lo SK, Priest N, Wake M. Socioeconomic Risk Factors for Mental Health Problems in 4-5-Year-Old Children: Australian Population Study. Acad Pediatr. 2010;

10. Huaqing qi C, Kaiser AP. Behavior Problems of Preschool Children From Low-Income Families: Review of the Literature. Topics Early Child Spec Educ. 2003;

11. Kessler RC, Berglund P, Demler O, Jin R, Merikangas KR, Walters EE. Lifetime prevalence and age-of-onset distributions of DSM-IV disorders in the national comorbidity survey replication. Archives of General Psychiatry. 2005.

12. Richards M, Abbott R. Childhood mental health and life chances in post-war Britain Insights from three national birth cohort studies. Centre for Mental Health. 2009.

13. Prince M, Patel V, Saxena S, Maj M, Maselko J, Phillips MR, et al. No health without mental health. Vol. 370, Lancet. 2007.

14. Costello EJ, Compton SN, Keeler G, Angold A. Relationships between Poverty and Psychopathology: A Natural Experiment. J Am Med Assoc. 2003;

15. Rehm J, Shield KD. Global Burden of Disease and the Impact of Mental and Addictive Disorders. Curr Psychiatry Rep. 2019 Jan 1;21(2):1–7.

16. Braveman P, Cubbin C, Daly MC, Duncan G, McDonough P, Williams DR. Optimal ses indicators cannot be prescribed across all outcomes (multiple letters). American Journal of Public Health. 2003.

17. Assis SG, Avanci JQ, Carvalhaes de Oliveira R de V. Socioeconomic inequalities and child mental health. Rev Saude Publica. 2009;

18. Pascoe JM, Wood DL, Duffee JH, Kuo A. Mediators and adverse effects of child poverty in the United States. Pediatrics. 2016;

19. Keister L. Income and wealth are not highly correlated: here is why and what it means [Internet]. Work In Progress. 2018 [cited 2020 Nov 9]. Available from: http://www.wipsociology.org/2018/10/29/income-and-wealth-are-not-highly-correlated-here-is-why-and-what-it-means/

20. Saez E, Zucman G. Wealth in equality in the United States since 1913: Evidence from capitalized income tax data. Q J Econ. 2016;

21. Killewald A, Pfeffer FT, Schachner JN. Wealth inequality and accumulation. Annual Review of Sociology. 2017.

22. Moulton V, Goodman A, Nasim B, Ploubidis GB, Gambaro L. Parental Wealth and Children’s Cognitive Ability, Mental, and Physical Health: Evidence From the UK Millennium Cohort Study. Child Dev. 2020;

23. Griffiths D, Lambert PS. Dimensions and boundaries: Comparative analysis of occupational structures using social network and social interaction distance analysis. Sociol Res Online. 2012;

24. Shavers VL. Measurement of socioeconomic status in health disparities research. J Natl Med Assoc. 2007;

25. Noble S, Mclennan D, Noble M, Plunkett E, Gutacker N, Silk M, et al. The English Indices of Deprivation 2019; Research report. Ministry of Housing, Communities and Local Government. 2019.

26. Visser K, Bolt G, Finkenauer C, Jonker M, Weinberg D, Stevens Gwjm. Neighbourhood deprivation effects on young people’s mental health and well-being: A systematic review of the literature. Social Science and Medicine. 2021.

27. Thapar A, McGuffin P. Validity of the shortened Mood and Feelings Questionnaire in a community sample of children and adolescents: A preliminary research note. Psychiatry Res. 1998;

28. Rescorla LA, Ginzburg S, Achenbach TM, Ivanova MY, Almqvist F, Begovac I, et al. Cross-Informant Agreement Between Parent-Reported and Adolescent Self-Reported Problems in 25 Societies. J Clin Child Adolesc Psychol. 2013;

29. Khanam R, Nghiem S, Rahman M. The income gradient and child mental health in Australia: does it vary by assessors? Eur J Heal Econ. 2020;

30. Johnston DW, Propper C, Pudney SE, Shields MA. The income gradient in childhood mental health: All in the eye of the beholder? J R Stat Soc Ser A Stat Soc. 2014;177(4).

31. Centre for Longitudinal Studies. Millenium Cohort Study Dataset [Internet]. UK Data Services. 2018. Available from: https://beta.ukdataservice.ac.uk/datacatalogue/series/series?id=2000031#!/abstract

32. Wampold BE, Serlin RC. The consequence of ignoring a nested factor on measures of effect size in analysis of variance. Psychol Methods. 2000;

33. Goodman R. The strengths and difficulties questionnaire: A research note. J Child Psychol Psychiatry Allied Discip. 1997;

34. Youth In Mind. Scoring the SDQ [Internet]. 2016 [cited 2020 Nov 12]. Available from: https://www.sdqinfo.org/py/sdqinfo/c0.py

35. Vugteveen J, de Bildt A, Theunissen M, Reijneveld M, Timmerman M. Validity Aspects of the Strengths and Difficulties Questionnaire (SDQ) Adolescent Self-Report and Parent-Report Versions Among Dutch Adolescents. Assessment. 2019;

36. Angold A, Costello J, Van Kämmen W, Stouthamer-Loeber M. Development of a short questionnaire for use in epidemiological studies of depression in children and adolescents: factor composition and structure across development. Int J Methods Psychiatr Res. 1996;

37. Thabrew H, Stasiak K, Bavin LM, Frampton C, Merry S. Validation of the Mood and Feelings Questionnaire (MFQ) and Short Mood and Feelings Questionnaire (SMFQ) in New Zealand help-seeking adolescents. Int J Methods Psychiatr Res. 2018;

38. Kessler RC, Andrews G, Colpe LJ, Mroczek DK. Short screening scales to monitor population prevalences and trends in non-specific psychological distress. 2002 [cited 2021 Apr 15]; Available from: https://www.researchgate.net/publication/11174017

39. Rubin D. Multiple imputation for nonresponse in surveys [Internet]. 2004 [cited 2021 Mar 3]. Available from: https://books.google.co.uk/books?hl=en&lr=&id=bQBtw6rx_mUC&oi=fnd&pg=PR24&ots=8PoJdP-XeN&sig=l35sET7zBnhtKf9-sLUTfAwNz5U

40. Little RJA, Rubin DB. Statistical Analysis with Missing Data, 3rd Edition | Wiley. 2019 [cited 2022 Apr 29];464. Available from: https://www.wiley.com/en-us/Statistical+Analysis+with+Missing+Data%2C+3rd+Edition-p-9780470526798

41. StataCorp. Stata Statistical Software: Release 16. College Station, TX: StataCorp LLC.; 2019.

42. Burnham KP, Anderson DR. Model selection and multimodel inference. 2002;208.

43. Hutton K, Nyholm M, Nygren JM, Svedberg P. Self-rated mental health and socio-economic background: A study of adolescents in Sweden. BMC Public Health [Internet]. 2014 Apr 23 [cited 2021 Mar 3];14(1):394. Available from: http://bmcpublichealth.biomedcentral.com/articles/10.1186/1471-2458-14-394

44. Thompson EJ, Richards M, Ploubidis GB, Fonagy P, Patalay P. Changes in the adult consequences of adolescent mental ill-health: Findings from the 1958 and 1970 British birth cohorts. Psychol Med. 2021;

45. Patalay P, Fitzsimons E. Development and predictors of mental ill-health and wellbeing from childhood to adolescence. Soc Psychiatry Psychiatr Epidemiol. 2018;

46. Collishaw S, Furzer E, Thapar AK, Sellers R. Brief report: a comparison of child mental health inequalities in three UK population cohorts. Eur Child Adolesc Psychiatry [Internet]. 2019 Nov 1 [cited 2021 Mar 22];28(11):1547–9. Available from: https://doi.org/10.1007/s00787-019-01305-9

47. Case A, Lubotsky D, Paxson C. Economic Status and Health in Childhood: The Origins of the Gradient. 2002.

48. Iqbal Z, Airey ND, Brown SR, Wright NMJ, Miklova D, Nielsen V, et al. Waiting list eradication in secondary care psychology: Addressing a National Health Service blind spot. Clin Psychol Psychother [Internet]. 2021 Jan 11 [cited 2021 Mar 24];cpp.2551. Available from: https://onlinelibrary.wiley.com/doi/10.1002/cpp.2551

49. Conger RD, Ge X, Elder GH, Lorenz FO, Simons RL. Economic Stress, Coercive Family Process, and Developmental Problems of Adolescents. Child Dev. 1994;

50. Masarik AS, Conger RD. Stress and child development: A review of the Family Stress Model. Current Opinion in Psychology. 2017.

51. Wickham S, Whitehead M, Taylor-Robinson D, Barr B. The effect of a transition into poverty on child and maternal mental health: a longitudinal analysis of the UK Millennium Cohort Study. Lancet Public Heal. 2017;2(3):141–8.

52. Conger RD, Conger KJ, Martin MJ. Socioeconomic Status, Family Processes, and Individual Development. J Marriage Fam. 2010 Jun;72(3):685–704.

53. Bøe T, Øverland S, Lundervold AJ, Hysing M. Socioeconomic status and children’s mental health: Results: from the Bergen Child Study. Soc Psychiatry Psychiatr Epidemiol [Internet]. 2012 Oct 20 [cited 2021 Mar 22];47(10):1557–66. Available from: http://helse.uni.no/barnibergen

54. McLaughlin KA, Breslau J, Green JG, Lakoma MD, Sampson NA, Zaslavsky AM, et al. Childhood socio-economic status and the onset, persistence, and severity of DSM-IV mental disorders in a US national sample. Soc Sci Med [Internet]. 2011 Oct [cited 2020 Nov 10];73(7):1088–96. Available from: https://pubmed.ncbi.nlm.nih.gov/21820781/

55. Albert D, Chein J, Steinberg L. The Teenage Brain: Peer Influences on Adolescent Decision Making [Internet]. Vol. 22, Current Directions in Psychological Science. SAGE Publications Inc.; 2013 [cited 2021 Apr 16]. p. 114–20. Available from: http://journals.sagepub.com/doi/10.1177/0963721412471347

56. Lê-Scherban F, Brenner AB, Schoeni RF. Childhood family wealth and mental health in a national cohort of young adults. SSM - Popul Heal [Internet]. 2016 Dec 1 [cited 2020 Nov 10];2:798–806. Available from: http://dx.doi.org/10.1016/j.ssmph.2016.10.008

57. Cowell F, Karagiannaki E, McKnight A. The changing distribution of wealth in the pre-crisis US and UK: The role of socio-economic factors [Internet]. Vol. 71, Oxford Economic Papers. Oxford University Press; 2019 [cited 2021 Mar 23]. p. 1–24. Available from: https://academic.oup.com/oep/article/71/1/1/5139797

58. Machin S. Houses and schools: Valuation of school quality through the housing market. Labour Econ. 2011;

59. Elgar FJ, McKinnon B, Torsheim T, Schnohr CW, Mazur J, Cavallo F, et al. Patterns of Socioeconomic Inequality in Adolescent Health Differ According to the Measure of Socioeconomic Position. Soc Indic Res. 2016;

60. Galster G, Booza J. The Mechanism (s) of Neighborhood Effects. Theory, Evidence, and Policy Implications The Mechanism (s) of Neighborhood Effects. Springer [Internet]. 2010;(January):33. Available from: https://link.springer.com/chapter/10.1007/978-94-007-2309-2_2

61. Brazil N, Clark WAV. Individual mental health, life course events and dynamic neighbourhood change during the transition to adulthood. Heal Place. 2017 May 1;45:99–109.

62. World Health Organization, Calouste Gulbenkian Foundation. WHO | Social determinants of mental health. WHO [Internet]. 2019 [cited 2021 Mar 24]; Available from: http://www.who.int/mental_health/publications/gulbenkian_paper_social_determinants_of_mental_health/en/

63. Riley AW. Evidence that school-age children can self-report on their health. Ambul Pediatr. 2004 Jul 1;4(4 SUPPL.):371–6.

64. Garbarski D. Perceived social position and health: Is there a reciprocal relationship? Soc Sci Med. 2010;

65. Ridley MW, Rao G, Schilbach F, Patel VH. Poverty, Depression, and Anxiety: Causal Evidence and Mechanisms. NBER Work Pap. 2020;

66. Sterne JAC, White IR, Carlin JB, Spratt M, Royston P, Kenward MG, et al. Multiple imputation for missing data in epidemiological and clinical research: potential and pitfalls. BMJ [Internet]. 2009 Jun 29 [cited 2022 Apr 29];338(7713):157–60. Available from: https://www.bmj.com/content/338/bmj.b2393

67. Pepinsky TB. A Note on Listwise Deletion versus Multiple Imputation. Polit Anal. 2018 Oct 1;26(4):480–8.

