## Supplementary file for "Socio-economic inequalities in adolescent mental health in the UK: multiple socio-economic indicators and reporter effects"

This PDF file contains all supporting materials for our manuscript. Section 1 contains descriptive statistics, including the differences between our cohort and analytical sample, SEP indicators missingness and collinearity tests for our five SEP indicators. The results from the drop one analyses can be found in section 2. Section 3 contains a model exploring the variance of parent internalising mental health explained by SEP indicators. Section 4 covers the results for the planned additional analyses on binary-level adolescent internalising symptoms and sex-SEP interaction.

**Table of contents**

Section 1- Descriptive statistics

*Table S1. Summary of means (95% confidence intervals), and proportions (%) for differences between the full cohort and analytic sample*

*Table S2. Missing values for SEP indicator variables*

*Table S3. Correlation matrix of SEP indicators*

*Table S4. Collinearity test of SEP indicators*

Section 2- Drop-one analysis

*Table S5. Log-likelihood tests using drop-one analysis of standardised adolescent mental health by SEP indicators at age 14*

Section 3- Parents mental health

*Figure S1. Variance of parents self-reported and parent adolescent reported internalising mental health explained by SEP indicators at age 14*

Section 4- Planned additional analysis

*Figure S2. Visualisation of clinical levels of adolescent internalising mental ill health's association with SEP indicators at age 14*

*Figure S3a. Visualisation of the interaction between sex and SEP indicators and their association with parent-reported adolescent internalising mental health at age 14*

*Figure S3b. Visualisation of the interaction between sex and SEP indicators and their association with adolescent self-reported internalising mental health at age 14*

**Section 1- Descriptive statistics**

*Table S1. Summary of means (95% confidence intervals), and proportions (%) for differences between the full cohort and analytic sample*

| | | |
|---|---|---|
| ***Mental health*** | | |
| Age 14 self-reported internalising mental health | 5.65 [5.5;5.8] | 5.55 [5.44;5.67] |
| Age 14 parent-reported internalising mental health | 1.94 [1.89;1.99] | 2.03 [1.99;2.07] |
| Age 17 self-reported internalising mental health | 3.52 [3.46;3.58] | 3.49 [3.44;3.54 |
| Age 17 parent-reported internalising mental health | 1.97 [1.92;2.03] | 2.07 [2.02;2.12] |
| Parent self-reported internalising mental health | 10.14 [10.04;10.24] | 10.43 [10.35;10.51] |
| ***Sociodemographic variables*** | | |
| Gender (male) | 48.6 | 49.96 |
| ***Socio-economic variables*** | | |
| *Parent education* | | |
| NVQ5 (e.g. Undergraduate degree) | 10.15 | 9.45 |
| NVQ4 (e.g. Certificate of Higher Education, BTEC professional diploma) | 35.92 | 34.17 |
| NVQ3 (e.g. A levels, BTEC level 3 diploma) | 15.71 | 15.29 |
| NVQ2 (e.g. GCSEs grades A*-C, BTEC level 2 diploma) | 24.12 | 24.79 |
| NVQ1 (e.g. GCSEs grades D-G, BTEC level 1 diploma) | 5.96 | 6.4 |
| None of these | 8.11 | 9.88 |
| *Household income* | | |
| Quintile 5 (highest income) | 26.22 | 20 |
| Quintile 4 | 20.71 | 22.66 |
| Quintile 3 | 16.35 | 19.12 |
| Quintile 2 | 18.38 | 19 |
| Quintile 1 (lowest income) | 18.33 | 19.18 |
| *Total net household wealth* | 165064 [ 139316;190811] | 159173 [142905;175442] |
| *Occupational status* | | |
| NS-SEC Managerial and professional | 28.03 | 27.7 |
| NS-SEC Intermediate | 15.92 | 16.25 |
| NS-SEC Small employer | 2.06 | 6.22 |
| NS-SEC Low supervisory and technical | 6.26 | 2 |
| NS-SEC Semi-routine and routine | 17.61 | 15.93 |
| Not in work | 30.09 | 31.89 |
| *Relative neighbourhood deprivation* | | |
| Decile 10 (least deprived) | 9.68 | 10.8 |
| Decile 9 | 10.34 | 10.04 |
| Decile 8 | 9.49 | 8.64 |
| Decile 7 | 9.08 | 8.66 |
| Decile 6 | 9.52 | 9.4 |
| Decile 5 | 9.77 | 9.37 |
| Decile 4 | 9 | 9.45 |
| Decile 3 | 9.77 | 10.33 |
| Decile 2 | 11.38 | 11.45 |
| Decile 1 (most deprived) | 12.01 | 11.83 |

*Table S2. Missing values for SEP indicator variables*

| SEP indicator | | Value | Missing | % Missing |
|---|---|---|---|---|
| Parent education | | 10577 | 392 | 3.57 |
| Household income | | 7,534 | 3435 | 31.31 |
| Household wealth | Mortgage value | 9372 | 1579 | 14.4 |
| | House value | 9780 | 1189 | 10.84 |
| | Debt value | 9351 | 1618 | 14.75 |
| | Asset value | 7867 | 3102 | 28.28 |
| Parent occupation | | 10,686 | 283 | 2.58 |
| Relative neighbourhood deprivation | | 10964 | 5 | 0.05 |
| Total | | 76,131 | 11603 | 15.24083 |

*Table S3. Correlation matrix of SEP indicators*

| SEP indicators | 1 | 2 | 3 | 4 | 5 |
|---|---|---|---|---|---|
| 1. Parental education | 1.000 | 0.294 (p<0.001) | 0.272 (p<0.001) | 0.272 (p<0.001) | 0.361 (p<0.001) |
| 2. Household income | 0.294 (p<0.001) | 1.000 | 0.305 (p<0.001) | 0.289 (p<0.001) | 0.295 (p<0.001) |
| 3. Household wealth | 0.272 (p<0.001) | 0.305 (p<0.001) | 1.000 | 0.251 (p<0.001) | 0.337 (p<0.001) |
| 4. Occupational status | 0.272 (p<0.001) | 0.289 (p<0.001) | 0.251 (p<0.001) | 1.000 | 0.325 (p<0.001) |
| 5. Relative neighbourhood deprivation | 0.361 (p<0.001) | 0.295 (p<0.001) | 0.337 (p<0.001) | 0.325 (p<0.001) | 1.000 |

*Table S4. Collinearity test of SEP indicators*

| SEP indicators | Variance Inflation Factor |
|---|---|
| Parent education | 1.58 |
| Household income | 1.46 |
| Household wealth | 1.45 |
| Parent occupational status | 1.39 |
| Relative neighbourhood deprivation | 1.25 |

**Section 2- Drop-one analysis**

*Table S5. Log-likelihood tests using drop-one analysis of standardised adolescent mental health by SEP indicators at age 14*

| | Imputed data sets that variable adds a unique contribution to (%) | |
|---|---|---|
| SEP Indicator | Parent report | Adolescent report |
| Parental education | 100 | 100 |
| Household income | 100 | 100 |
| Household wealth | 100 | 100 |
| Occupational Status | 100 | 100 |
| Relative neighbourhood deprivation | 96 | 100 |

**Section 3- Parents mental health**

*Figure S1. Variance of parents self-reported and parent adolescent reported internalising mental health explained by SEP indicators at age 14*

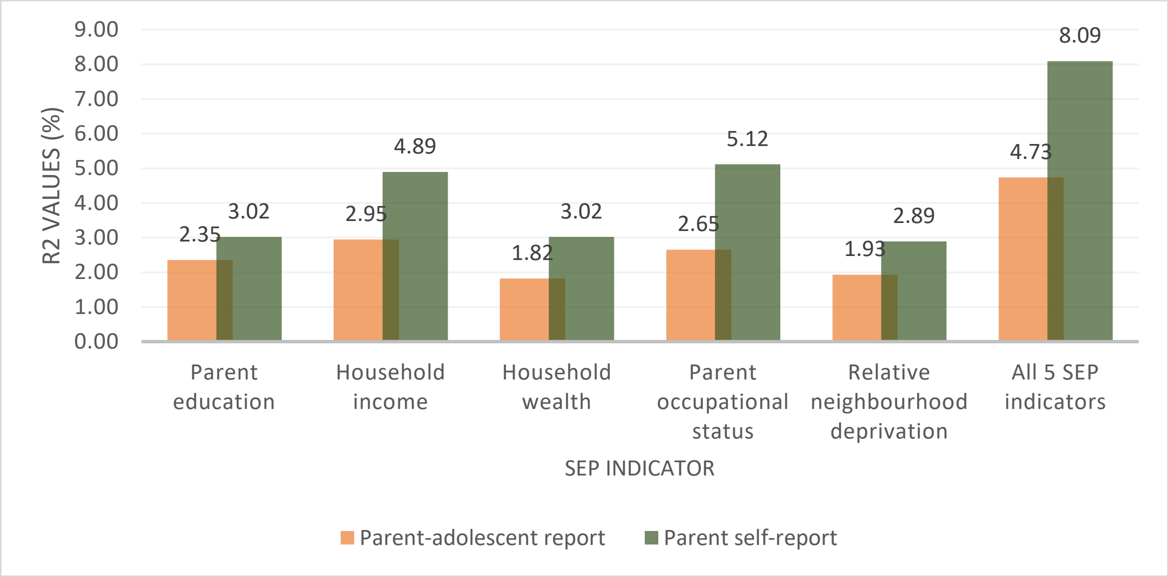

### Section 4- Planned additional analysis

*Figure S2. Visualisation of clinical levels of adolescent internalising mental ill health's association with SEP indicators at age 14*

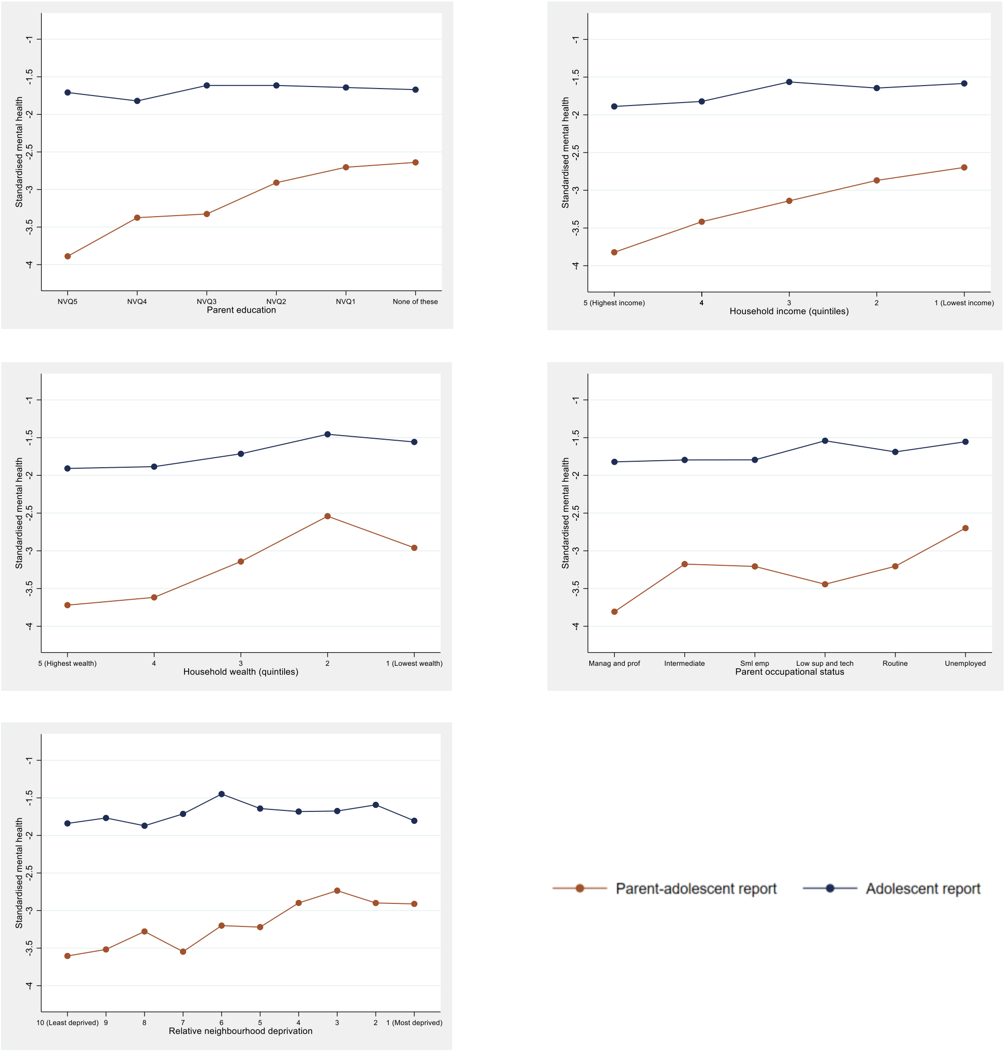

*Figure S3a. Visualisation of the interaction between sex and SEP indicators and their association with parent-reported adolescent internalising mental health at age 14*

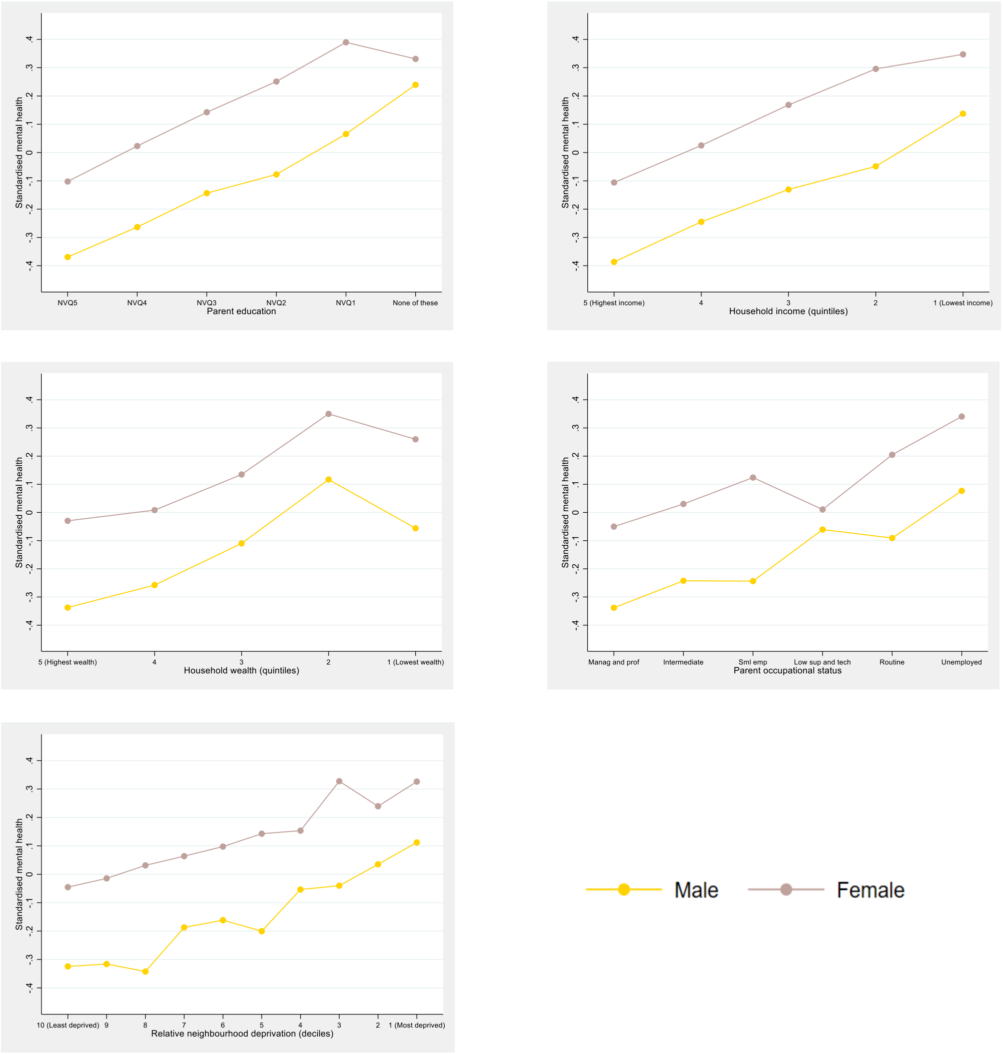

*Figure S3b. Visualisation of the interaction between sex and SEP indicators and their association with adolescent self-reported internalising mental health at age 14*

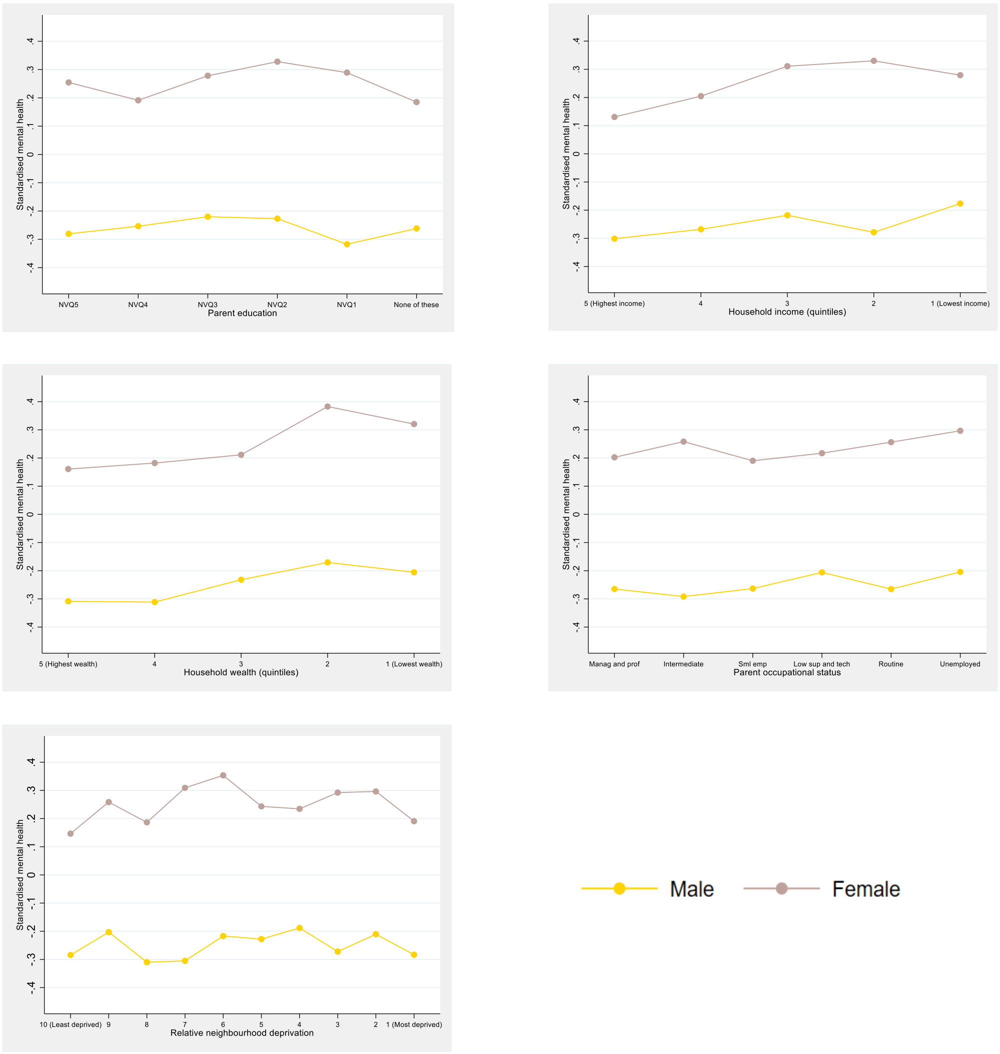
